# Structural Variation Detection and Association Analysis of Whole-Genome-Sequence Data from 16,905 Alzheimer’s Diseases Sequencing Project Subjects

**DOI:** 10.1101/2023.09.13.23295505

**Authors:** Hui Wang, Beth A Dombroski, Po-Liang Cheng, Albert Tucci, Ya-Qin Si, John J Farrell, Jung-Ying Tzeng, Yuk Yee Leung, John S Malamon, The Alzheimer’s Disease Sequencing Project, Li-San Wang, Badri N Vardarajan, Lindsay A Farrer, Gerard D Schellenberg, Wan-Ping Lee

## Abstract

Structural variations (SVs) are important contributors to the genetics of numerous human diseases. However, their role in Alzheimer’s disease (AD) remains largely unstudied due to challenges in accurately detecting SVs. Here, we analyzed whole-genome sequencing data from the Alzheimer’s Disease Sequencing Project (ADSP, N=16,905 subjects) and identified 400,234 (168,223 high-quality) SVs. We found a significant burden of deletions and duplications in AD cases (OR=1.05, *P*=0.03), particularly for singletons (OR=1.12, *P*=0.0002) and homozygous events (OR=1.10, *P*<0.0004). On AD genes, the ultra-rare SVs, including protein-altering SVs in *ABCA7*, *APP*, *PLCG2*, and *SORL1*, were associated with AD (SKAT-O *P*=0.004). Twenty-one SVs are in linkage disequilibrium (LD) with known AD-risk variants, e.g., a deletion (chr2:105731359-105736864) in complete LD (R^2^=0.99) with rs143080277 (chr2:105749599) in *NCK2*. We also identified 16 SVs associated with AD and 13 SVs associated with AD-related pathological/cognitive endophenotypes. Our findings demonstrate the broad impact of SVs on AD genetics.

**Search Terms:** Alzheimer’s disease, Structural variation, Copy number variation

## Introduction

Alzheimer’s disease (AD) is a neurodegenerative disease characterized by abnormal deposits of extracellular Aβ plaques and intracellular neurofibrillary tangles^1^. Typically, the accumulation of these neuropathological changes is accompanied by neuronal death, leading to various symptoms such as memory loss, apathy, difficulty swallowing, and walking^2^. Among individuals aged 65 and older, AD has an incidence rate of 10.7% and is the fifth-leading cause of death^2^.

Genetic factors play a significant role in the etiology of AD, with the estimate of heritability ranging from 58% to 79%^3^. However, genetic risk factors identified in previous studies explain only a limited portion of heritability in AD. Mutations in *APP*, *PSEN1*, and *PSEN2* cause an early-onset form of AD that is inherited as an autosomal dominant trait with high penetrance, but these mutations only account for about 11% of early-onset AD that is approximately 0.6% of all AD^4^. *APOE* genotype is the most prominent genetic risk factor for AD, and it is estimated that approximately 40-50% of individuals diagnosed with AD carry at least one copy of the *APOE* ε4 risk allele^5^. Overall, variations in *APP*, *PSEN1*, *PSEN2*, and *APOE* explain 20-50% of total genetic variance (heritability) of AD, with *APOE* ε4 accounting for most of this fraction due to its high frequency^6^.

In the past decade, genome-wide association studies (GWASs) identified > 75 additional AD risk loci^7–10^. However, compared to *APOE* alleles, variants at those loci have a small effect size or are rare in the population, contributing little to the overall heritability. *APOE* alleles alone can achieve an AUC (Area Under the Receiver Operating Characteristic Curve) of 0.70 in predicting AD, whereas the best AUC is only 0.61 when all other common single nucleotide variants (SNVs) are combined^11^. Even if all common SNVs, including *APOE* alleles, are considered, they only account for 24-33% of phenotypic variance^12,13^, which is much lower than the estimated heritability of AD and thus suggests a role for other genetic mechanism.

Structural variants (SVs) are genomic alterations larger than 50bp that include deletions, duplications, inversions, insertions, translocations, and complex combinations of these events. SVs contribute more to individual genetic variation in terms of total nucleotide content, and thus the difference in genomic sequences between two humans can increase from 0.1% with SNVs alone to 1.5% when SVs are considered^14^. Moreover, SVs can have profound effects on diseases and other traits by disrupting gene function and regulation or modifying gene dosage through copy number changes, deleting exons, and creating new splicing acceptors or donor sites. Therefore, analyzing SVs has the potential to identify new genetic associations and account for the missing heritability in AD.

SVs have been identified in several genes implicated in AD. For instance, duplications in *APP* have been found to be the causal factor for autosomal dominant early-onset AD in a few families^15–19^. In addition, a deletion in exon 9 of *PSEN1* was identified in families with a form of early-onset AD characterized by spastic paraparesis and atypical plaques^20,21^. A low-copy repeat of 18 Kb in length within *CR1*, which creates an additional C3b/C4b-binding site, may account for some GWAS signals in the *CR1* region^22,23^. The 1 Mb region on 17q21.31 containing *MAPT* has two major haplotypes H1 and H2, which are characterized by a ∼900[Kb inversion flanked by a few duplication blocks and tagged by a 238 bp deletion between exons 9 and 10 of *MAPT*^24^. The H1 and H2 haplotypes are associated with a range of neurodegenerative diseases including AD^24–26^. Additionally, copy number variants (CNVs) in *AMY1*, which are correlated with salivary amylase protein level and digestion of starchy food, are associated with AD. Individuals with high copy numbers (≧10) of *AMY1* have a significantly lower risk of developing AD^27^. These examples show that identification and analysis of SVs in AD genetics hold great potential for uncovering new genetic associations and providing a more comprehensive understanding of the genetic underpinnings of this complex disease.

To discover SV variants possibly contributing to AD risk, we evaluated SVs detected in whole-genome sequence (WGS) data from 16,905 subjects from the Alzheimer’s Disease Sequencing Project (ADSP). We detected 400,234 SVs and found rare SVs in known AD genes, including *SORL1*, *ABCA7* and *APP,* as well as SVs in linkage disequilibrium (LD) with AD GWAS signals. Moreover, we found an increased burden of deletions and duplications (particularly, singleton and homozygous events) in AD and identified possible risk SVs in *ADD3*, *ITPR2*, and *NTM* through association analysis.

## Results

### SV discovery and characteristics

The SV discovery pipeline, including the Manta^28^, Smoove^29^, Svimmer^30^, and GraphTyper2^30^ SV callers (**Methods**), was applied to ADSP^31^ R3 release (NG00067.v7) WGS data (N=16,905; **Table 1**). We observed 400,234 SVs (231,385 deletions, 45,839 duplications, 119,648 insertions, and 3,362 inversions) of which 168,223 (98,805 deletions, 24,602 duplications, 44,130 insertions, and 506 inversions) were classified as high-quality calls (**Table S1**, **Methods**). Notably, genotype calls for deletions exhibited superior quality with a lower missing genotype rate compared to duplications, insertions, and inversions (**Fig. S1**). This observation highlights the higher quality of deletion detection on WGS over other SV types using available callers.

**Table 1.** Characteristics of study participants (N = 16,905)

|  | **AD**<br>**(N = 6,646)** | **Control**<br>**(N = 6,938)** | **Unknown**<br>**AD status**<br>**(N = 3,321)** |
| --- | --- | --- | --- |
| **Age (SD)** | 74.62 (10.44) | 77.40 (7.98) | 72.04 (9.28) |
| **Sex (%)** |  |  |  |
| Female | 3,998 (60%) | 4,639 (67%) | 1,586 (48%) |
| Male | 2,648 (40%) | 2,299 (23%) | 1,735 (52%) |
| **APOE status<sup>a</sup> (%)** |  |  |  |
| ε4 | 3,552 (54%) | 2,188 (32%) | 972 (30%) |
| No ε4 | 3,084 (46%) | 4,661 (68%) | 2,279 (70%) |
| **Ancestry<sup>b</sup> (%)** |  |  |  |
| European | 4,381 (66%) | 3,214 (46%) | 2,871 (86%) |
| African American | 1,454 (22%) | 2,036 (29%) | 129 (4%) |
| Latin American | 769 (12%) | 1,655 (24%) | 253 (8%) |
| East Asian | 18 (0.27%) | 9 (0.13%) | 32 (0.96%) |
| Other | 24 (0.36%) | 24 (0.35%) | 36 (1%) |
| **Ethnicity (%)** |  |  |  |
| non-Hispanic | 5,523 (85%) | 4,912 (71%) | 1,058 (80%) |
| Hispanic | 1,022 (15%) | 2,003 (29%) | 265 (20%) |

AD, Alzheimer's disease; Age, age at onset for individuals with AD or age at last exam for non-AD subjects; SD, standard deviation.
<sup>a</sup>APOE ε4 status is based on rs429358 observed from whole genome sequencing data.
<sup>b</sup>Ancestry is inferred using GRAF-pop<sup>37</sup>.

On average, each individual had 14,607 (3,875 high-quality) deletions, 764 (288 high-quality) duplications, 6,980 (2,504 high-quality) insertions, and 19 (3 high-quality) inversions. Individuals of African ancestry had more SV calls compared to individuals of other ancestries (**Fig. 1A**), possibly because the human reference genome is biased towards European ancestry or higher level of genetic diversity in Africans^32–34^. Similar to SNVs, the first two principal components of common SVs distinguished samples from different ancestral backgrounds (**Fig. 1B**). However, the third principal component of SVs is associated with read length and sequencing platforms (**Fig. S2**), indicating batch effect is an important confounding factor to consider when performing SV analysis.

**Fig. 1:**
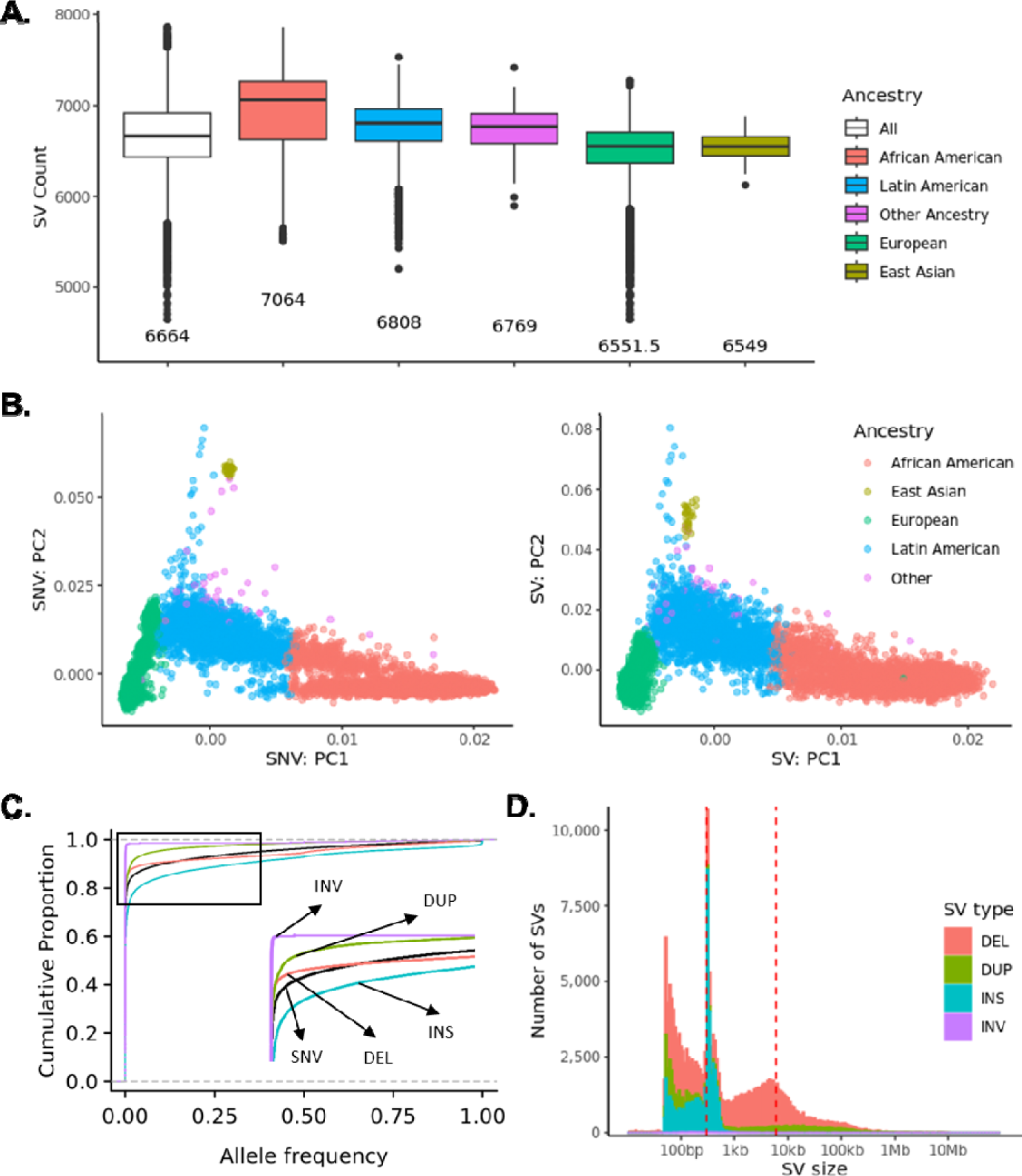
Characteristics of high-quality SVs. **A.** Number of high-quality SVs per individual by ancestry. **B.** Principal component analysis of high-quality SV with MAF > 0.01 and Hardy-Weinberg Equilibrium (HWE) > 1e-5. **C.** The cumulative fractions of variants by allele frequency. **D.** The size distribution of high-quality SVs.

Comparable to the allele frequency (AF) distribution of SNVs, most SVs are extremely rare. Among 400,234 SVs, 94,923 (24%) are singletons, and 232,295 (58%) are rare with AF < 1% (**Fig. S3**). When considering the 168,223 high-quality SVs, 67,595 (40%) are singletons, and 140,164 (83%) are rare with AF < 1%. **Fig. 1C** shows that the AF distribution of deletions is more similar to the AF distribution of SNVs compared to other SV types. Analysis of the size of the SVs revealed two peaks centered around 300 bp and 6,000 bp (**Fig. 1D**), suggesting the possibility that many SVs are introduced by transposons, particularly, Alus (∼300 bp) and L1s (∼6,000 bp).

Functional annotation analysis performed using AnnotSV^35^ showed that rare SVs are more likely to be deleterious than common SVs (Wilcoxon Rank Sum *P* < 0.0001) (**Fig. 2A**). This finding was confirmed using annotation from VEP^36^, which shows that protein-altering SVs tend to be rare (odds ratio [OR] = 4.71, Chi-Square *P* < 0.0001, **Fig. 2B**). Additionally, we observed a higher proportion of singletons SVs in coding or regulatory regions (**Fig. 2C**), suggesting negative selection against deleterious SVs in functionally important regions of the genome. Overall, our results highlight the importance of evaluating rare SVs when studying genetic variation in human disease.

**Fig. 2:**
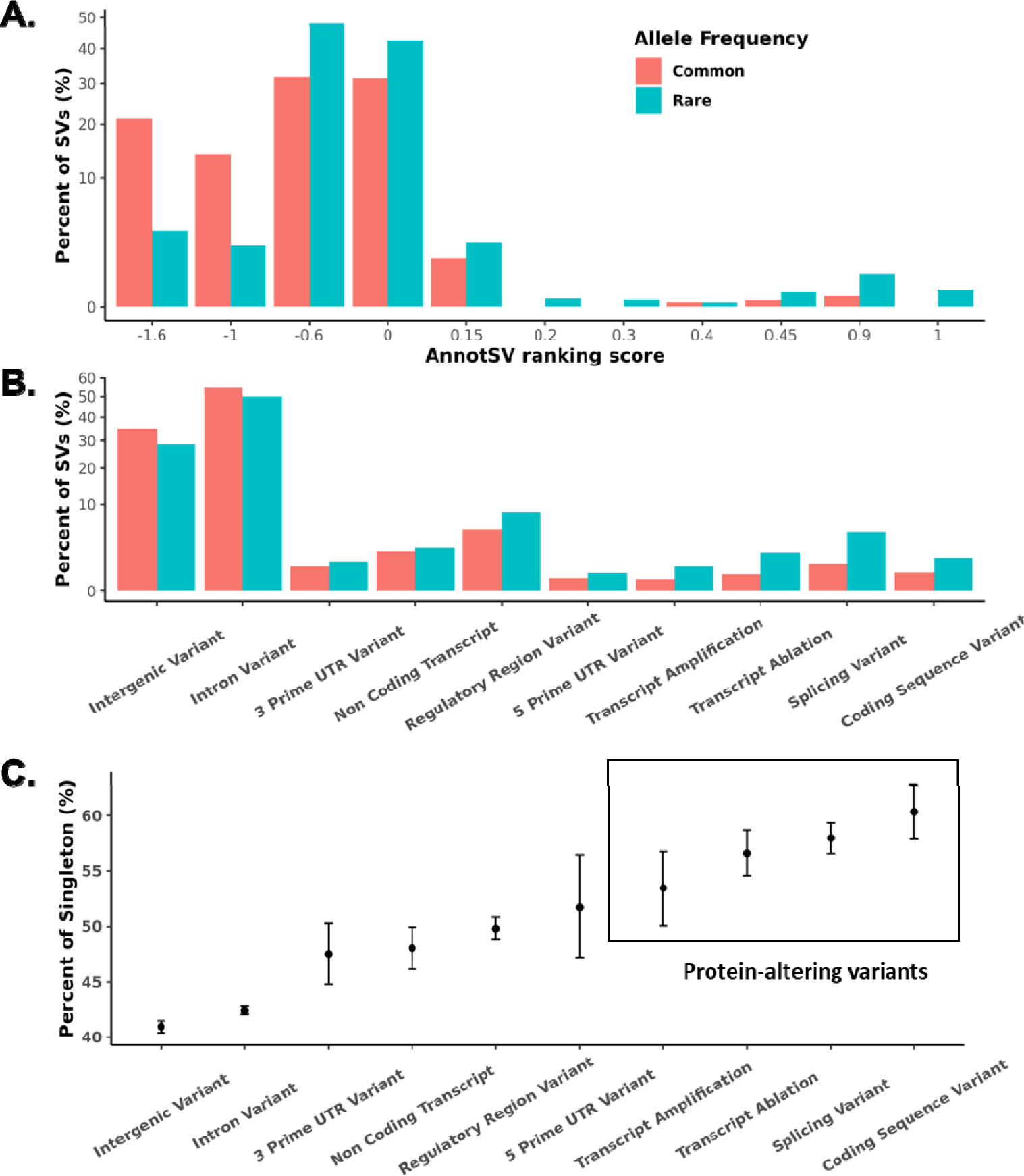
Functional annotation of SVs. **A.** AnnotSV ranking scores of common (AF ≥ 0.01) and rare (AF < 0.01) high-quality SVs. The rare SVs are more likely to be deleterious with higher AnnotSV ranking scores. **B.** VEP annotation of common (AF ≥ 0.01) and rare (AF < 0.01) high-quality SVs. The protein-altering SVs tend to be rare. **C.** Percent of singletons in a specified functional category by VEP.

### SV quality evaluation and laboratory validation

Evaluation of the sensitivity of SV calling pipeline using synthetic mutations^38^ (**Methods**) revealed a sensitivity of 99.4% for 4,000 deletions and 94.4% for 1,500 inversions (**Table S2**). We did not perform an evaluation for insertions since the inserted sequences and positions are ambiguous in the simulation of synthetic mutations.

Then, we evaluated our SV call set against external SV databases. Approximately 50% of the high-quality SVs were detected in the Genome Aggregation Database (gnomAD, 292,307 SV sites), but there was less overlap with SVs in the 1000 Genomes Project (1KG, 66,505 SV sites) (**Fig. S4**). The difference was due to fewer samples in 1KG compared to gnomAD. The SV callset before high-quality filtering had a higher recall (a higher percentage of SVs from gnomAD and 1KG) at the cost of lower precision (a lower percentage of SVs confirmed by gnomAD and 1KG) (**Fig. S4, Fig. S5**).

Of 95 SVs selected for experimental validation (**Table S3**; **Methods**), 78 were confirmed, resulting in a sensitivity rate of 82%. When considering only high-quality SVs, the sensitivity increased to 85% with 61 out of 72 SVs being experimentally validated. On individual genotype level, an accuracy of 89% was achieved for 276 called genotypes for 95 SVs undergoing PCR validation (**Table S4**), and this value increased to 92% for 207 called genotypes for 72 high-quality SVs (**Table S4**).

### SVs in linkage disequilibrium with known AD risk loci

SVs are larger genomic perturbations and may have more severe functional impact compared to SNVs; therefore, SVs in LD with AD GWAS risk SNVs are more likely to account for the statistical association in the regions, especially if the SNVs are not predicted to have an impact on protein structure or gene expression. We identified 21 SVs (12 deletions, two duplications, and seven insertions) that are in LD with established AD GWAS loci^8–10^ (**Table 2**). Three deletions, in particular, showed high LD (R^2^ > 0.9) with GWAS signals near or in *NCK2*, *NBEAL1*, and *TMEM106B*. A 5.5 Kb deletion (chr2:105731359-105736864) located 8 Kb upstream of *NCK2* is in perfect LD (R^2^ = 0.99) with rs143080277 (chr2:105749599), which is a rare variant (AF = 0.005) in the intron of *NCK2*^10^. A 5.2 Kb deletion (chr2:203034369-203039560) in *NBEAL1* intron and overlapping with H3K27ac peak from Encode^39^ is in high LD (R^2^ = 0.94) with rs139643391 (chr2:202878716)^10^, which is a 3 prime UTR variant of *WDR12*. A 323 bp (chr7:12242077-12242399) Alu deletion located on the exon 8 of *TMEM106B* is in LD with *TMEM106B* intronic variants, rs5011436 (chr7:12229132, R^2^ = 0.92)^9^ and rs13237518 (chr7:12229967, R^2^ = 0.90)^10^, which are not only associated with the risk of AD but also protect carriers of *C9ORF72* repeat expansion from the risk of frontotemporal dementia^40^.

**Table 2.**
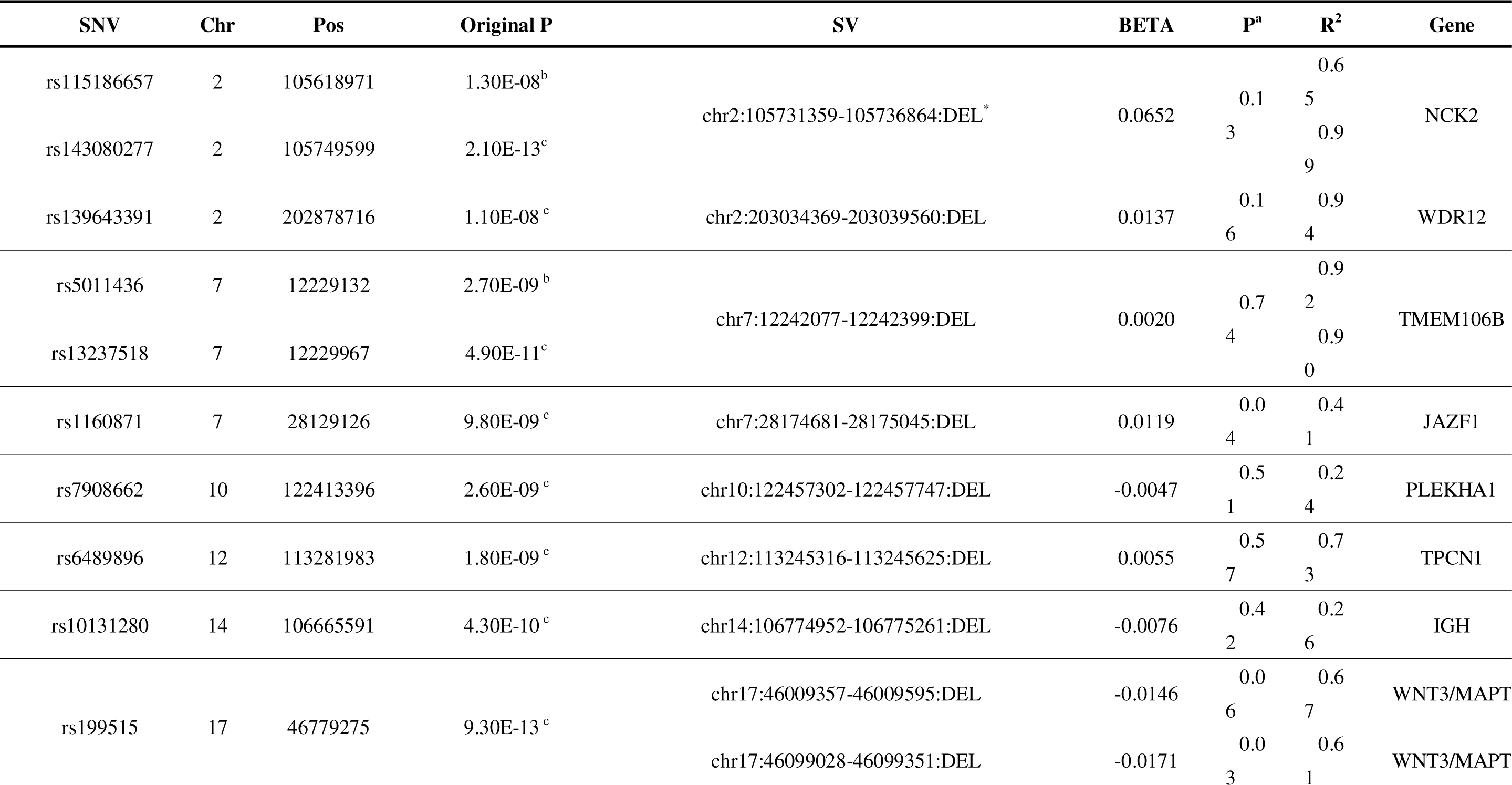
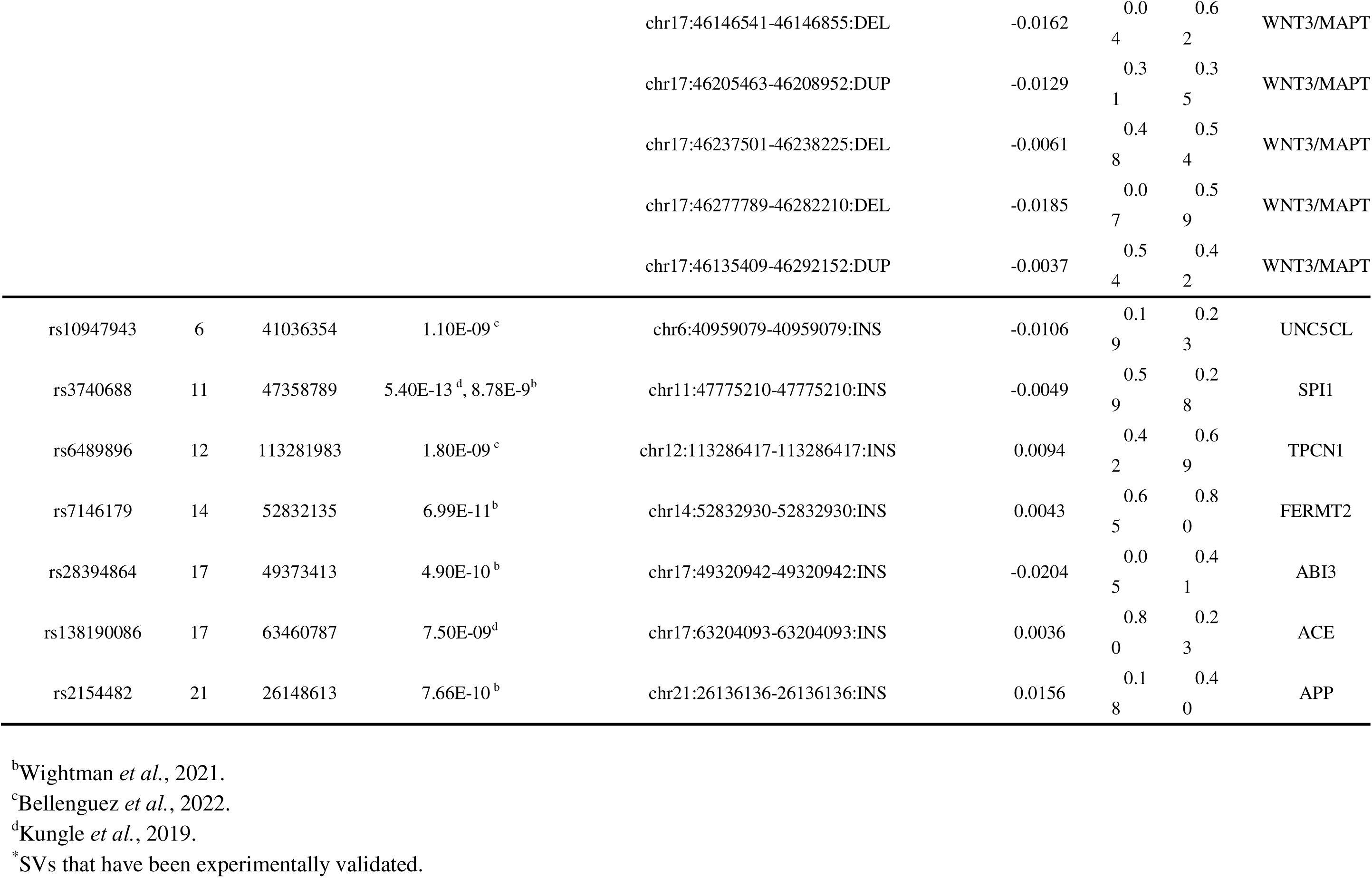
High-confident SVs in linkage disequilibrium with AD GWAS signals.

Other deletions that are in moderate LD (0.2 < R^2^ < 0.9) with GWAS signals can impact exons, enhancers, transposons, and conserved regions (**Table 2**). A 446 bp deletion (chr10:122457302-122457747) extending into exon 2 of *ARMS2* is in LD (R^2^ = 0.24) with rs7908662 (chr10:122413396, *PLEKHA1* intronic variant)^10^. A 310 bp deletion (chr14:106774952-106775261) overlaps an enhancer element in the IGH gene cluster and is in LD (R^2^ = 0.26) with rs10131280 (chr14:106665591)^10^. A 310 bp Alu deletion (chr12:113245316-113245625) in *TPCN1* intron is in LD (R^2^ = 0.73) with rs6489896 (chr12:113281983)^10^. A 365 bp deletion (chr7:28174681-28175045) in *JAZF1* intron overlapping evolutionally conserved sequence defined by phastCons and phyloP is in LD (R^2^ = 0.41) with rs1160871 (chr7:28129126)^10^.

The *MAPT* H1/H2 haplotype, defined by a 900 kb inversion and tagged by numerous SNVs, has been associated with several neurodegenerative diseases, including progress supranuclear palsy, frontotemporal disorders, Parkinson’s disease, and AD^24,25,41^. We identified five deletions and two duplications in moderate LD (R^2^ = 0.35-0.67, **Table 2**) with a H1/H2 tagging SNV (rs199515, chr17:46779275), which is associated with AD^10^. These SVs further confirmed the complex genomic structure in the region and highlight the difficulty in identifying the causal variants within the H1/H2 haplotype. **Table 2** also describes seven high-quality insertions, excluding those in problematic regions (**Methods**), that are in LD with AD GWAS signals.

### SVs on AD risk/protective genes

We first focused on SVs that were reported to be associated with AD in previous studies^43–47^. Ten rare SVs (**Table S5**) were replicated in our SV callset. A 417 Kb duplication (**Fig. S6**) covering the *APP* is identified in one individual with early onset of AD at his age of 52. Subsequently, we noticed two other carriers of duplication who were dropped from the initial analysis due to failed quality control. One individual having the duplication was his sibling and developed AD at age of 49, and the other individual is his sibling’s offspring who developed AD at age of 53. This finding provides compelling evidence that the duplication of *APP* is a rare cause of autosomal dominant early-onset AD^15–17,19^. A 7.68 Mb inversion covering the entire 21q21.2 is identified in one individual with early onset of AD at her age of 60 years old. The inversion was experimentally validated, and the alignments showed clear breakpoints of the inversion (**Fig. S7**). In addition, the 5.6 Kb deletion, covering exons 2-5 of *HLA-DRA* found in nine AD cases by Swaminathan *et al.*^43^, are present in eight samples in our analysis, including five AD cases (three showed early onset of AD with age < 65) and three unclear-AD-status individuals (two are diagnosed as progressive supranuclear palsy, and the remaining one is with BRAAK stage 2). A few other SVs, encompassing *GBE1*, *EPHA5*, and *EVC*, are replicated in our dataset.

SVs on AD risk/protective genes could interfere with protein function and lead to disease. Therefore, we identified 77 high-confident SVs (**Methods**), including 44 deletions, 15 duplications, and 18 insertions on AD risk/protective genes determined by the ADSP gene verification committee (see Table2 on https://adsp.niagads.org/gvc-top-hits-list/). Nine deletions and five duplications have an allele count ≥ 5 (AF ranging from 0.0002 to 0.4690), but none of them were significantly associated with AD (**Table S6**), and none of these SVs were tagged known AD-associated SNVs. The remaining 35 deletions and 10 duplications are ultra-rare (MAC < 5), of which 34 (25 deletions and 9 duplications) are singletons (**Table 3**). We performed an aggregated analysis of 45 ultra-rare CNVs (35 deletions and 10 duplications), using SKAT-O test^48^ instead of calculating individual p-values given the limited statistical power due to low allele count, and observed a significant association with AD status (*P* = 0.0050), highlighting the contribution of ultra-rare CNVs to the etiology of AD.

**Table 3.** Ultra-rare SVs on AD genes.

| SV | Size | AC (Case)<br>[AgeOnset,Sex,Eth] | AC (Control)<br>[Age,Sex,Eth] | Gene | Type | Protein-altering<sup>a</sup> |
| --- | --- | --- | --- | --- | --- | --- |
| chr19:1050368-1050973<sup>b</sup> | 605 | 4<br>[74, F, AA]<br>[69, M, AA]<br>[81, F, AA]<br>[81, F, AA] | 0 | ABCA7 | DEL | Yes |
| chr16:81775821-81829769<sup>b</sup> | 53,948 | 4<br>[75, M, E]<br>[56, F, E]<br>[-, M, E]<br>[-, M, E] | 0 | PLCG2 | DEL | Yes |
| chr19:1052156-1060559 | 8,403 | 3<br>[72, F, AA]<sup>d</sup><br>[72, F, AA] | 1<br>[70, F, L] | ABCA7 | DUP | Yes |
| chr16:81860089-81940500 | 80,411 | 3<br>[89, F, E]<br>[56, F, E]<br>[-, F, E] | 0 | PLCG2 | DEL | Yes |
| chr2:127094683-127094740<sup>b</sup> | 57 | 2<br>[86, M, E]<br>[87, F, E] | 1<br>[84, F, E] | BIN1 | DEL | No |
| chr7:100296733-100385675 | 88,942 | 2<br>[85, F, E]<br>[56, F, E] | 0 | PILRA | DEL | Yes |
| chr16:81907252-81907401 | 149 | 2<br>[67, F, E]<br>[61, F, E] | 0 | PLCG2 | DEL | No |
| chr14:92611452-92611515 | 63 | 2<br>[90, M, E]<br>[61, F, E] | 0 | RIN3 | DEL | No |
| chr19:1043504-1053484 | 9,980 | 1 [78, M, E] | 0 | ABCA7 | DEL | Yes |
| chr19:1054326-1061615<sup>b</sup> | 7,289 | 1 [67, M, E] | 0 | ABCA7 | DUP | Yes |
| chr15:58720431-58721649 | 1,218 | 1 [79, F, AA] | 0 | ADAM10 | DEL | No |
| chr21:25815144-26232105<sup>e</sup> | 416,961 | 1 [52, M, E] | 0 | APP | DUP | Yes |
| chr21:25958556-25971275 | 12,719 | 1 [-, F, E] | 0 | APP | DEL | No |
| chr21:26163874-26163976 | 102 | 1 [70, M, E] | 0 | APP | DUP | No |
| chr2:127102503-127104954 | 2,451 | 1 [70, M, AA] | 0 | BIN1 | DEL | No |
| chr10:113725274-113726288 | 1,014 | 1 [75, F, E] | 0 | CASP7 | DEL | Yes |
| chr11:60138757-60178011 | 39,254 | 1 [65, F, AA] | 1 [82, F, E] | MS4A6A | DEL | Yes |
| chr11:60179765-60450406<sup>b</sup> | 270,641 | 1 [-, F, O] | 0 | MS4A6A | DUP | Yes |
| chr11:86050643-86054032 | 3,389 | 1 [70, F, AA] | 0 | PICALM | DEL | No |
| **chr16:81734058-81749307<sup>c</sup>** | 15,249 | 1 [-, F, E] | 0 | PLCG2 | DEL | Yes |
| chr16:81749081-81749132 | 51 | 1 [75, M, AA] | 0 | PLCG2 | DEL | No |
| **chr16:81755550-81764402<sup>c</sup>** | 8,852 | 1 [84, F, E] | 0 | PLCG2 | DEL | Yes |
| chr16:81772599-81777744 | 5,145 | 1 [69, F, E] | 0 | PLCG2 | DEL | Yes |
| chr16:81792449-81792587 | 138 | 1 [68, F, E] | 0 | PLCG2 | DEL | No |
| chr16:81798030-81802305<sup>b</sup> | 4,275 | 1 [72, M, E] | 1 [89, F, AA] | PLCG2 | DEL | Yes |
| chr16:81822810-81822862<sup>b</sup> | 52 | 1 [83, M, E] | 0 | PLCG2 | DEL | No |
| chr16:81868226-81868350 | 124 | 1 [66, F, AA] | 1 [70, M, AA] | PLCG2 | DEL | No |
| chr14:92369331-92644481 | 275,150 | 1 [85, M, AA] | 0 | RIN3 | DUP | Yes |
| chr14:92531510-92573409 | 41,899 | 1 [-, F, E] | 0 | RIN3 | DUP | Yes |
| chr11:121303351-121495669 | 192,318 | 1 [61, F, AA] | 0 | SORL1 | DUP | Yes |
| chr11:121490959-121499413 | 8,454 | 1 [70, F, E] | 0 | SORL1 | DEL | Yes |
| chr19:1051380-1051420 | 40 | 0 | 1 [81, M, AA] | ABCA7 | DEL | No |
| chr15:58621731-58622007 | 276 | 0 | 1 [64, F, L] | ADAM10 | DUP | No |
| chr19:44909364-44909819 | 455 | 0 | 1 [80, M, AA] | APOE | DUP | Yes |
| chr21:26013065-26013159 | 94 | 0 | 1 [69, F, AA] | APP | DEL | No |
| chr2:127064222-127064288 | 66 | 0 | 1 [89, F, AA] | BIN1 | DEL | No |
| chr2:127092466-127092526 | 60 | 0 | 1 [68, M, L] | BIN1 | DEL | No |
| chr2:127094016-127102983 | 8,967 | 0 | 1 [80, F, E] | BIN1 | DEL | No |
| chr1:207604022-207605343 | 1,321 | 0 | 1 [-, F, L] | CR1 | DEL | No |
| chr7:100377717-100378235 | 518 | 0 | 1 [64, M, AA] | PILRA | DEL | No |
| chr16:81746327-81746435<sup>b</sup> | 108 | 0 | 1 [85, F, E] | PLCG2 | DEL | No |
| chr16:81885235-81893072 | 7,837 | 0 | 1 [73, F, AA] | PLCG2 | DEL | Yes |
| chr14:73215181-73313643 | 98,462 | 0 | 1 [84, F, E] | PSEN1 | DEL | Yes |
| chr14:92535796-92535871<sup>b</sup> | 75 | 0 | 1 [85, F, E] | RIN3 | DEL | No |
| chr14:92605335-92607867<sup>b</sup> | 2,532 | 0 | 1 [77, F, E] | RIN3 | DEL | No |

<sup>a</sup>SV overlaps with gene exons.
<sup>b</sup>SVs that are experimentally validated.
<sup>c</sup>SVs with read depth and split reads support but no PCR product for flanking primers.
<sup>d</sup>Represents homozygous even
<sup>e</sup>Two additional family individuals had the duplication but failed quality control due to less confident genotypes. Nevertheless, alignment evidence strongly supports the presence of duplication. Their onset ages are 49 and 53.
F, female; M, male; E, European, A, Asian; AA, African American; L, Latin American; O, other; “-” represents missing age. AD risk/protective genes are selected by ADSP Gene Verification Committee (https://adsp.niagads.org/gvc-top-hits-list/).

Notably, 14 of the 35 ultra-rare deletions and 8 of the 10 ultra-rare duplications are protein altering variants. For instance, we identified in *SORL1* a 192 Kb duplication spanning exons 1-5 and an 8 Kb deletion affecting exon 6 (**Fig. 4**). Previous studies indicated that *SORL1* deficiency can lead to AD through defects in the endolysosome-autophagy network^49,50^, and nearly all individuals with damaging SNVs in *SORL1* developed AD^51^. Eight out of nine individuals with *ABCA7* exonic deletions or duplications in our data (**Fig. S8**) developed AD, supporting previous studies that observed loss-of-function *ABCA7* variants among AD cases^52^. We also found protein-altering ultra-rare deletions and duplications in *APP*, *PLCG2*, *PILRA*, *CASP7*, *MS4A6A*, *RIN3*, *APOE*, and *PSEN1* (**Table 3**). In particular, 17 of 21 individuals with ultra-rare deletions in *PLCG2* were AD cases (SKAT-O *P* = 0.029). We also identified 18 high-quality insertions located in AD genes (**Table S7**). However, the aggregated effect of these insertions on AD risk was not significant (SKAT-O *P* = 0.21).

**Fig. 4:**
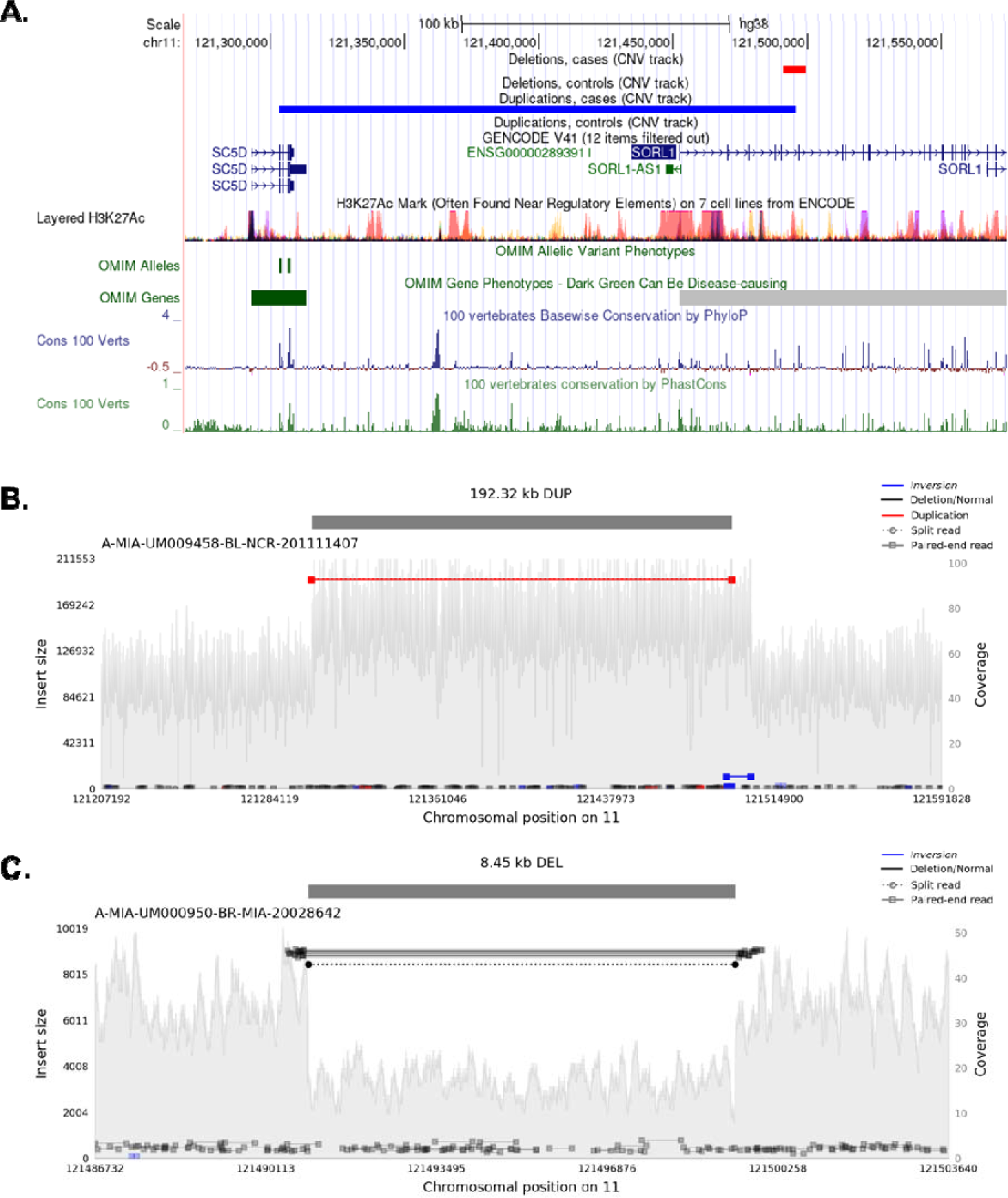
Ultra-rare deletion and duplication on *SORL1*. **A.** Deletion and duplication on *SORL1*. **B.** The 192 Kb duplication covers part of *SORL1* and *SC5D*. **C.** The 8.45 Kb deletion covers exon 6 of *SORL1*.

### SV burden in AD

We performed burden tests of SVs, including CNVs (deletions and duplications), insertions, and inversions separately and collectively, and found a moderate burden of CNVs in AD cases (OR = 1.05, *P* = 0.0321), but no significant burden of insertions and inversions was detected (**Table S8**). The increased CNV burden in AD cases was driven by the presence of singletons (OR = 1.12, *P* = 0.0002) and homozygous CNVs (OR = 1.10, *P* = 0.0004). This is consistent with the burden of ultra-rare CNVs in AD genes, in which 34 out of 45 ultra-rare CNVs are singletons. The result suggests that singletons and homozygous CNVs, which were not considered in previous association analyses, may be important contributors to the genetic basis of AD.

### SVs associated with AD and AD endophenotypes

From our association analysis using 12,908 subjects (6,328 AD cases and 6,580 controls, excluding subjects with unknown AD diagnosis and SV quality outliers, **Methods**), six common and nine rare SVs were found associated with AD at a false discovery rate (FDR) < 0.2 (**Table 4**, **Fig. 5A**). Notably, a 12.7 Kb (chr10:110025269-110037941, AF = 0.000426) deletion in the intron of *ADD3* was exclusively found in 11 AD cases and not in any control. In gnomAD, this deletion has a lower AF of 0.000277, which may be attributed to fewer AD cases in gnomAD. Moreover, there is a rare SNV (rs773892439) in complete LD (R^2^ = 1) with this deletion. Since the SNV is extremely rare (gnomAD AF of 0.00022, TOPMed^53^ AF of 0.00033, and our AF of 0.00065), it was not included in previous GWASs. Another rare deletion (chr12:26731939-26732033, AF = 0.00155) in *ITPR2* was found in 33 AD cases and 7 controls. The deletion is in intron 2 of *ITPR2*, which may be a regulatory region as indicated by the H3K4me1 and H3K27ac signals as well as transcription factor ChIP-seq clusters in this region (**Fig. S11A**). *ITPR2* was found to be widely expressed across different brain regions (**Fig. S11B**), with a higher expression in AD (**Fig. S11C**). SVs in *LMNTD1*, *LHFPL6*, *RNA5SP293*, *RABGAP1*, *ADD3*, *ITPR2*, and *CLIC4* were confirmed by PCR validation.

**Fig. 5:**
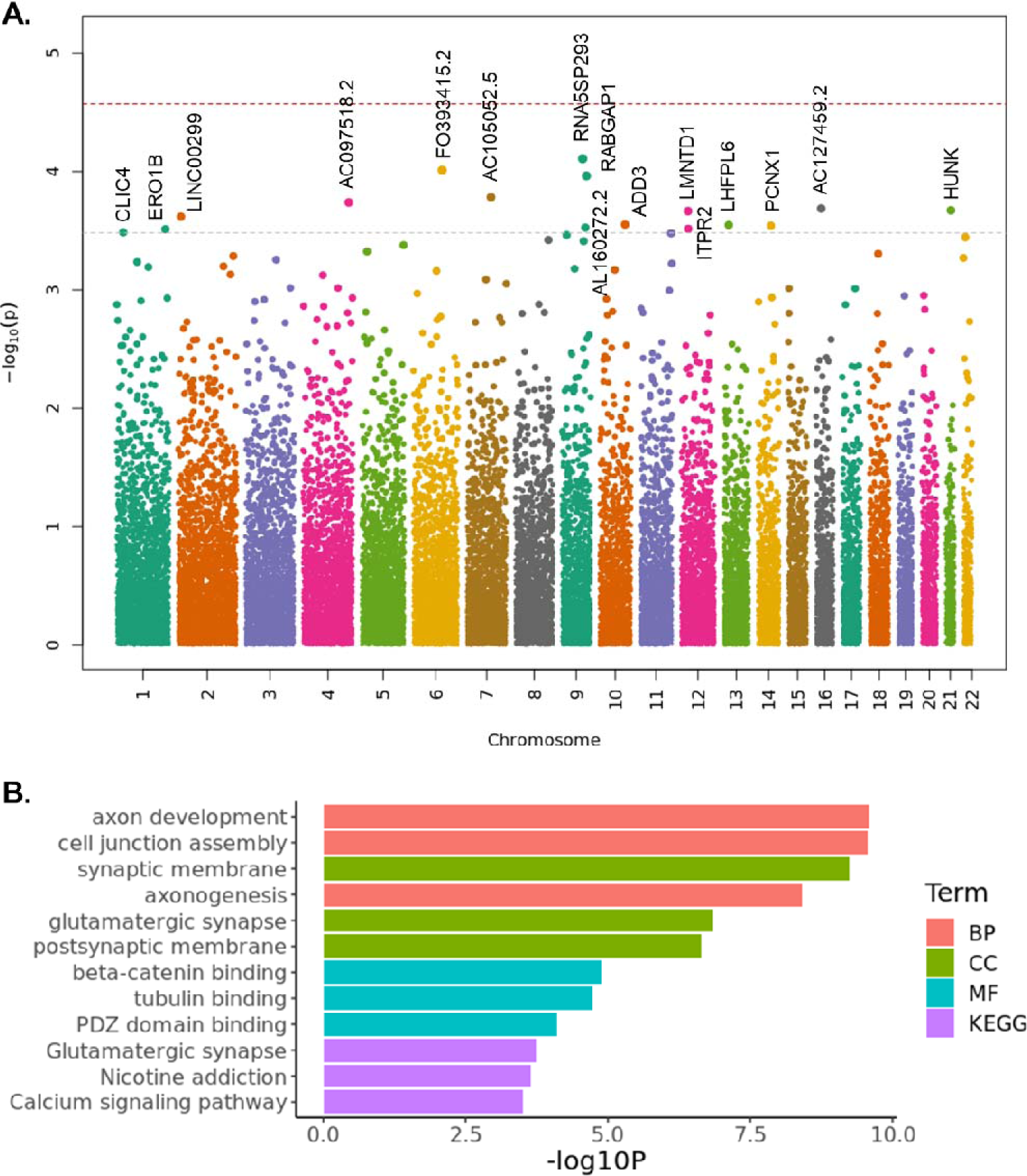
Association of SVs with AD and enrichment analysis. **A.** Association of SVs with AD. Red line represents an FDR of 0.05. Gray line represents an FDR of 0.2. **B.** Enrichment analysis for high-quality SVs (nominal *P* < 0.05) that are not in problematic regions. BP, Biological Process; CC, Cellular Component; MF, Molecular Function; KEGG, Kyoto Encyclopedia of Genes and Genomes.

**Table 4.**
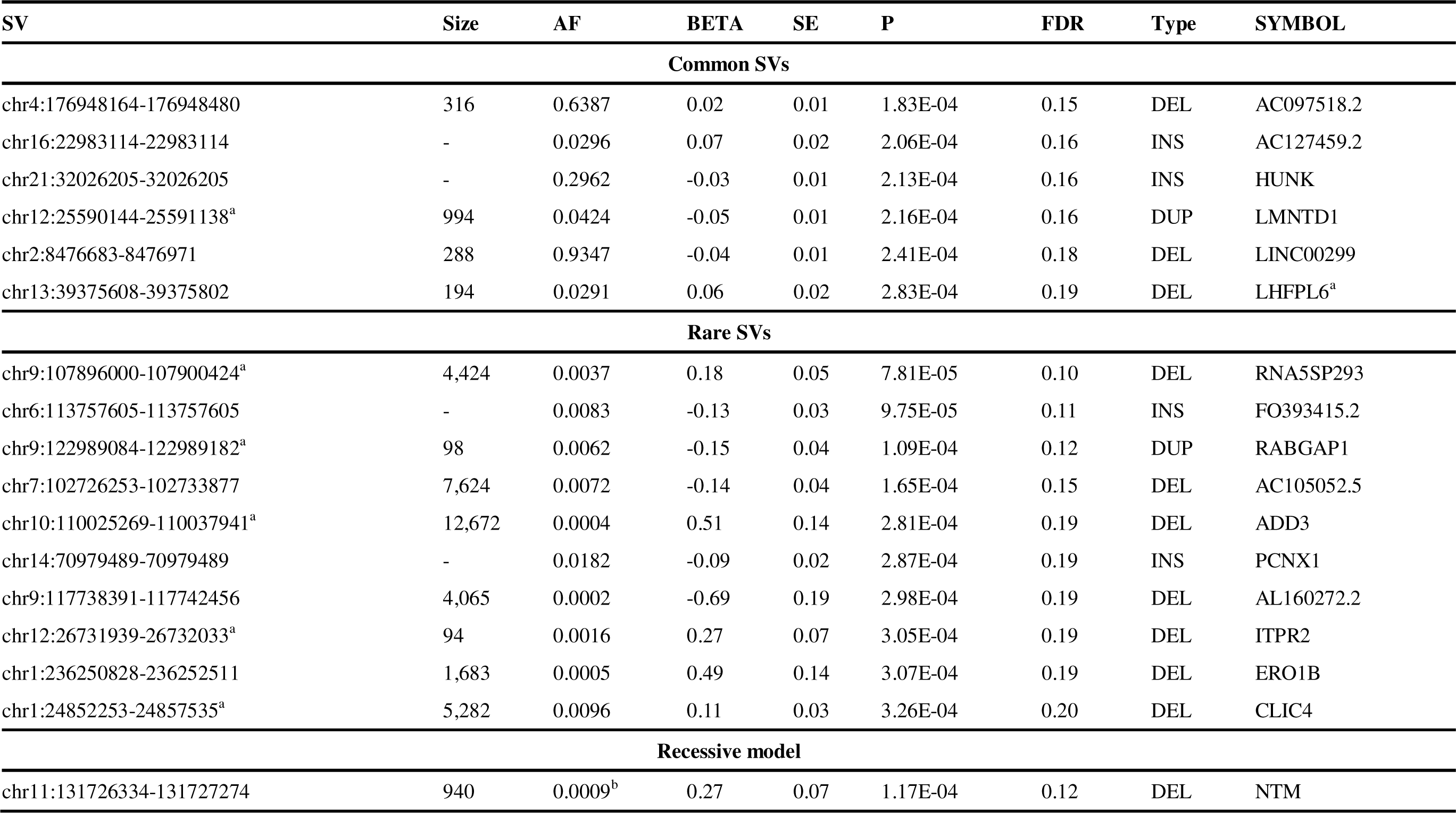
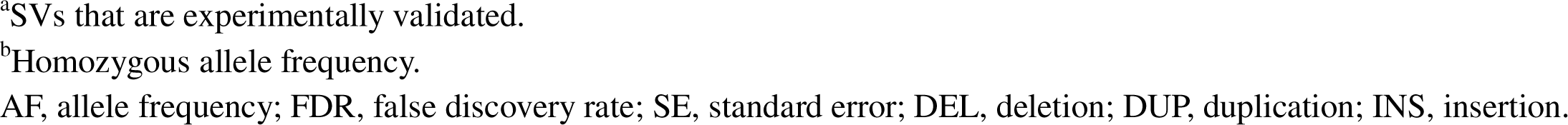
Association analysis of AD status (FDR < 0.2)

| SV | Size | AF | BETA | SE | P | FDR | Type | SYMBOL |
| --- | --- | --- | --- | --- | --- | --- | --- | --- |
| **Common SVs** |  |  |  |  |  |  |  |  |
| chr4:176948164-176948480 | 316 | 0.6387 | 0.02 | 0.01 | 1.83E-04 | 0.15 | DEL | AC097518.2 |
| chr16:22983114-22983114 | - | 0.0296 | 0.07 | 0.02 | 2.06E-04 | 0.16 | INS | AC127459.2 |
| chr21:32026205-32026205 | - | 0.2962 | -0.03 | 0.01 | 2.13E-04 | 0.16 | INS | HUNK |
| chr12:25590144-25591138<sup>a</sup> | 994 | 0.0424 | -0.05 | 0.01 | 2.16E-04 | 0.16 | DUP | LMNTD1 |
| chr2:8476683-8476971 | 288 | 0.9347 | -0.04 | 0.01 | 2.41E-04 | 0.18 | DEL | LINC00299 |
| chr13:39375608-39375802 | 194 | 0.0291 | 0.06 | 0.02 | 2.83E-04 | 0.19 | DEL | LHFPL6<sup>a</sup> |
| **Rare SVs** |  |  |  |  |  |  |  |  |
| chr9:107896000-107900424<sup>a</sup> | 4,424 | 0.0037 | 0.18 | 0.05 | 7.81E-05 | 0.10 | DEL | RNA5SP293 |
| chr6:113757605-113757605 | - | 0.0083 | -0.13 | 0.03 | 9.75E-05 | 0.11 | INS | FO393415.2 |
| chr9:122989084-122989182<sup>a</sup> | 98 | 0.0062 | -0.15 | 0.04 | 1.09E-04 | 0.12 | DUP | RABGAP1 |
| chr7:102726253-102733877 | 7,624 | 0.0072 | -0.14 | 0.04 | 1.65E-04 | 0.15 | DEL | AC105052.5 |
| chr10:110025269-110037941<sup>a</sup> | 12,672 | 0.0004 | 0.51 | 0.14 | 2.81E-04 | 0.19 | DEL | ADD3 |
| chr14:70979489-70979489 | - | 0.0182 | -0.09 | 0.02 | 2.87E-04 | 0.19 | INS | PCNX1 |
| chr9:117738391-117742456 | 4,065 | 0.0002 | -0.69 | 0.19 | 2.98E-04 | 0.19 | DEL | AL160272.2 |
| chr12:26731939-26732033<sup>a</sup> | 94 | 0.0016 | 0.27 | 0.07 | 3.05E-04 | 0.19 | DEL | ITPR2 |
| chr1:236250828-236252511 | 1,683 | 0.0005 | 0.49 | 0.14 | 3.07E-04 | 0.19 | DEL | ERO1B |
| chr1:24852253-24857535<sup>a</sup> | 5,282 | 0.0096 | 0.11 | 0.03 | 3.26E-04 | 0.20 | DEL | CLIC4 |
| **Recessive model** |  |  |  |  |  |  |  |  |
| chr11:131726334-131727274 | 940 | 0.0009<sup>b</sup> | 0.27 | 0.07 | 1.17E-04 | 0.12 | DEL | NTM |

<sup>a</sup>SVs that are experimentally validated.
<sup>b</sup>Homozygous allele frequency.
AF, allele frequency; FDR, false discovery rate; SE, standard error; DEL, deletion; DUP, duplication; INS, insertion.

Under a nominal P < 0.05, there are 2,411 high-quality SVs not in the problematic regions (**Methods**). Enrichment analysis of the 2,411 SVs revealed an over-representation of neuronal function-related terms, such as axon development and synaptic membrane (**Fig. 5B**). Among the 2,411 SVs, 37 are protein-altering variants (**Table S9**), including protein-altering variants in genes that have been found to be related to AD, e.g., *NTN3* and *CIB2*^54,55^.

Since a significant homozygous CNV burden is detected, we performed association using a recessive model, of which assumes that two copies of the alternative allele are required to alter the risk. As a result, a 1 Kb deletion (chr11:131726334-131727274) in the intron of *NTM* is the only SV with FDR < 0.2 using the recessive model. Interestingly, the variants inside *NTM* have been associated with tau pathology in previous studies ^56,57^.

In addition, we extended our association analysis to endophenotypes. **Table 5** shows six common and six rare SVs with an FDR < 0.2 for cognitive functions, CSF biomarkers, and neuropathologic measurements. No significant genomic inflation was observed for all endophenotypes (**Fig. S12**), indicating that confounding factors are well adjusted. The most significant signal is a rare deletion (chr4:188173309-188183202, AF = 0.0028, *P* = 1.72 × 10^-08^) located in the intergenic region that is a transcription factor binding site. A rare SNV (rs1418703978) which shows even lower AF (gnomAD AF of 0.00019, TOPMed AF of 0.00026, and our AF of 0.00047) is in complete LD with the deletion. A 100 Kb deletion (chr6:31391686-31488609) encompassing the entire *MICA* gene is associated with amyloid presence (*P* = 1.09 × 10^-07^). Previous studies showed that the *MICA* deletion is accompanied by a *MICB* null allele (MICB0107N)^58^, indicating loss of function of both *MICA* and *MICB*. These genes are located in the MHC locus, which has been found associated with AD risk^59^.

**Table 5.** Association analysis of AD endophenotypes (FDR < 0.2)

| Phenotype | SV | Size | AF | BETA | SE | P | FDR | Type | SYMBOL |
| --- | --- | --- | --- | --- | --- | --- | --- | --- | --- |
| **Common SVs** |  |  |  |  |  |  |  |  |  |
| EXF | chr11:22412330-22412446 | 116 | 0.1800 | -0.10 | 0.02 | 2.93E-06 | 0.014 | DEL | SLC17A6 |
| MEM | chr8:1096443-1097561 | 1,118 | 0.0480 | -0.24 | 0.05 | 7.43E-06 | 0.041 | DEL | DLGAP2 |
| LAN | chr17:46810995-46811289 | 294 | 0.0968 | 0.15 | 0.03 | 2.02E-05 | 0.121 | DEL | WNT3 |
| EXF | chr1:4228390-4228390 | - | 0.1090 | -0.11 | 0.03 | 8.23E-05 | 0.136 | INS | EEF1DP6 |
| MEM | chr5:119121855-119122904 | 1,049 | 0.0416 | -0.25 | 0.06 | 3.79E-05 | 0.140 | DEL | DMXL1 |
| MEM | chr5:34754523-34754523 | - | 0.0986 | -0.14 | 0.03 | 4.99E-05 | 0.179 | INS | RAI14 |
| **Rare SVs** |  |  |  |  |  |  |  |  |  |
| REAG | chr4:188173309-188183202 | 9,893 | 0.0028 | -2.23 | 0.40 | 1.72E-08 | 0.001 | DEL | LINC02434 |
| AMY | chr6:31391686-31488609 | 96,923 | 0.0017 | -0.61 | 0.12 | 1.09E-07 | 0.010 | DEL | MICA |
| EXF | chr9:85662873-85662873 | - | 0.0051 | -0.58 | 0.12 | 3.57E-06 | 0.014 | INS | AGTPBP1 |
| EXF | chr18:48162022-48162075 | 53 | 0.0006 | -1.44 | 0.36 | 6.96E-05 | 0.119 | DUP | ZBTB7C |
| AMY | chr9:117738391-117742456 | 4,065 | 0.0011 | -0.66 | 0.15 | 6.39E-06 | 0.181 | DEL | AL160272.2 |
| LAN | chr3:33035060-33035060 | - | 0.0027 | -0.74 | 0.18 | 4.46E-05 | 0.195 | INS | GLB1 |

AF, allele frequency; FDR, false discovery rate; SE, standard error; DEL, deletion; DUP, duplication; INS, insertion; REAG, NIA-Reagan diagnosis of AD; AMY, amyloid presence (dichotomous); EXF, executive function score; MEM, memory score; LAN, language score.

## Discussion

The complexity of generating high-quality SVs on WGS for SV association analysis is challenging, and a major concern is to ensure the analysis is not based on false positive SVs. To achieve this, we developed a pipeline to filter SVs and employed stringent criteria during the burden analysis to only include high-quality SVs. For each significant SV, we examined read coverages and other alignment signals by Samplot and performed experimental validations if samples are available in the lab (**Methods**). Despite our efforts, false positive/negative calls on individual samples can still occur, which may undermine the result of the analysis. Therefore, we suggest a broader validation of significant SVs using long reads as the cost and accuracy of long reads improve rapidly.

We reported SVs in LD with known AD risk loci (such as SNVs in *NCK2*, *WDR12*, and *TMEM106B*) and on AD risk/protective genes (such as *APP*, *SORL1*, and *ABCA7*). Other than that, researchers can use our SV calling set to explore SVs on a particular gene of interest. For example, there are SVs on genes that might be related to the risk of disease by interacting with well-known AD genes (such as *PSEN2* and *APOE*). A deletion (chr1:226827423-226834076, near *PSEN2*) spanning the entire *lnc-PSEN2-7* and overlapping with a possible enhancer supported by H3K27ac signals (**Fig. S9**) was identified in an individual (Latin American ancestry, inferred by GRAF-pop^37^), who had onset of AD symptoms at age 71 years old. We also observed in one AD case an exonic deletion in *MPO* (**Fig. S10**), a gene that has been reported to affect AD risk through interact with *APOE*^60^.

Our association analysis yielded some interesting findings. One notable discovery is a 12 Kb deletion in *ADD3*, which is a gene encoding a subunit of adducin protein called γ-adducin and was reported associated with neural function. The α-adducin encoded by *ADD1* can either dimerize with β-adducin (*ADD2*) or γ-adducin (*ADD3*) to form the adducin protein^61^. Heterodimers of α-adducin and β-adducin are mainly in red blood cells and neurons as the expression of adducin β were tissue-specific and α-adducin and γ-adducin were present in most tissue types^61^. Adducin plays an essential role in the membrane cytoskeleton of red blood cells^62^ and is highly expressed in dendritic spines^63^ and growth cones of neurons^64^. Moreover, overexpression of γ-adducin promotes neurite-like process in COS7 cells^65^, suggesting important roles of adducin in brain function. Variants in *ADD3* were found to be associated with hypertension, cerebral palsy, renal disease, vascular disease and cognitive dysfunction^66,67^. Along with tau and a few other CDK5 substrates, γ-adducin is also hyperphosphorylated (possibly by CDK5) in APP/PS1 mice^68^. Interestingly, ADD3 displayed a significantly lower expression in 6-month-old APP/PS1 mice while significantly higher expression in 14-month-old APP/PS1 mice^69^. In addition, γ-adducin is in involved in trans-Golgi-network through re-organization of the actin network around the Golgi complex^65^, therefore, may be able to regulate intracellular trafficking of APP and relevant secretases.

Our study provided a valuable resource for the analysis of SVs in AD. We identified SVs from WGS data across a large cohort of AD participants with diverse ancestry. We reported SVs tagging AD risk SNVs, providing new mechanism of actions for GWAS signals. Deleterious rare SVs on well-known AD genes have been discovered. We found a higher burden of ultra-rare SVs on AD genes, and overall, higher burden of homozygous and singleton CNVs in AD patients. Finally, our association analysis revealed a few potential candidate SVs and genes that are worthy of further study.

## Methods

### Study subjects

Alzheimer’s Disease Sequencing Project (ADSP)^31^ is a collaborative project aiming at identifying new variants, genes, and therapeutic targets in AD. In the R3 release of ADSP, 16,905 subjects were collected across 24 cohorts and whole genome sequencing was performed by Illumina HiSeqX, HiSeq2000, HiSeq2500, and NovaSeq platforms. The ancestry of each individual was inferred using GRAF-pop^37^. The samples came from diverse ancestries with 10,466 Europeans, 3,619 African Americans, 2,677 Latin Americans, 59 East Asians, 84 of other ancestries. There are 6,646 AD cases, 6,938 controls and 3,321 subjects with unknown status in this study. Sample characteristics were displayed in **Table 1**.

After removing duplicates and subjects without AD diagnosis, 13,371 samples were kept for analysis. Then, 463 outlier subjects, with too many (> median + 4*MAD) SV calls or too few (< median - 4*MAD) high-quality SV calls, were removed (**Fig. S13**). There were 12,908 samples (6,328 cases and 6,580 controls) remaining for association analysis (**Fig. S14**). Compared to the samples that were kept for further analysis, outliers are more likely to be of smaller insert size and lower coverage (**Fig. S15**).

### SV calling

**Fig. S16** illustrates the SV calling pipeline. For each sample, SVs were called by Manta^28^ (v1.6.0) and Smoove^29^ (v0.2.5) with default parameters. Calls from Manta and Smoove were merged by Svimmer^30^ to generate a union of two call sets for a sample. Unresolved non-reference ‘breakends’ (BNDs) and SVs > 10 Mb were filtered. Then, all individual sample VCF files were merged together by Svimmer as input to Graphtyper2 (v2.7.3)^30^ for joint genotyping. SV calls after joint-genotyping are comparable across the samples, therefore, can be used directly in genome-wide association analysis^30^. The pipeline is available on https://github.com/whtop/SV-ADSP-Pipeline.

### SV selection by algorithmic models

Graphtyper2 annotates each SV call by algorithmic models, i.e., breakpoint, coverage, and aggregated models^30^. Note that an SV call can be annotated by multiple models so there will be duplicated records in VCF if an SV call has more than one algorithmic model. Aggregated model has the highest recall than the other two models^30^. Therefore, SVs were selected based on the order of aggregated, breakpoint, and then coverage models (**Table S1**).

### High-quality SVs

A subset of SV calls was defined as high-quality calls. The criteria for high-quality SVs can be found in Graphtyper2 study^30^: For deletion, QD (QUAL divided by non-reference sequence depth) > 12 & (ABHet (allele balance for heterozygous calls (read count of call2/(call1 + call2)) where the called genotype is call1/call2, −1 if no heterozygous calls.) > 0.30 | ABHet < 0) & (AC / NUM_MERGED_SVS (number of SVs merged)) < 25 & PASS_AC (number of alternate alleles in called genotyped that have “FT” field as “PASS”) > 0 & PASS_ratio (ratio of genotype calls that have “FT” field as “PASS”) > 0.1; For duplication, QD > 5 & PASS_AC > 0 & (AC / NUM_MERGED_SVS) < 25; For insertion, PASS_AC > 0 & (AC / NUM_MERGED_SVS) < 25 & PASS_ratio > 0.1 & (ABHet > 0.25 | ABHet < 0) & MaxAAS (maximum alternative allele support per alternative allele) > 4; For inversions: PASS_AC > 0 & (AC / NUM_MERGED_SVS) < 25 & PASS_ratio > 0.1 & (ABHet > 0.25 | ABHet < 0) & MaxAAS > 4. Then, if an SV still has multiple records in VCF due to multiple algorithmic models, we selected based on the order of aggregated, breakpoint, and then coverage models.

### Problematic regions

There are regions in the human genome that tend to have anomalous, or high signal in WGS experiments^70^. SVs that reside in those regions can be unreliable and should be reported. Specifically, we compiled problematic regions in the genome from the following sources: (1) the ENCODE blacklist: a comprehensive set of regions that could result in erroneous signal^71^; (2) the 1000 Genome masks: regions of the genome that are more or less accessible to next generation sequencing methods using short reads; (3) the set of assembly gaps defined by UCSC; (4) the set of segmental duplications defined by UCUC; (5) the low-complexity regions, satellite sequences and simple repeats defined by RepeatMasker (Tarailo-Graovac and Chen 2009).

### High-confident SVs

For any SVs reported on AD risk/protective genes and from association, Samplot^73^ was used to check their alignment supports of read depth and/or split reads if SV types are deletions, duplications, and inversions. For insertions, which cannot be inspected using Samplot, we kept insertions that are high-quality and not in the problematic regions.

### SV annotation

SVs were annotated using VEP (V 107)^36^ and annotSV (V 3.1.1)^35^. SVs that were annotated (by VEP) to be able to cause transcript ablation/amplification, stop gain, start/stop lost, frameshift, inframe deletion/insertion, missense mutation, and affecting splice acceptor/donor were classified as protein-altering variants. The impact of SVs is also evaluated by annotSV ranking score, which is an adaptation of the work provided by the joint consensus recommendation of the American College of Medical Genetics and Genomics (ACMG) and ClinGen^74^.

### SV validation

Structural variants from the 1000 Genomes Project phase III^75^ and gnomAD^76^ were downloaded from dbVar database^77^ with study accession ID estd219 and nstd166. On chromosomes 1-22, there are 66,505 and 292,307 SVs from the 1000 Genomes Project and gnomAD, respectively. For deletions/duplications/inversions, calls with at least 50% reciprocal overlapping were considered as replicated. For insertions, we searched for calls with breakpoints within 500bp. Then, we estimated sensitivity of Graphtyper2 by synthetic mutations (i.e., “spiking-in” SVs) generated from three samples by Malamon *et al.*^38^.

For PCR validation, the sequence surrounding the variants was extracted and used to design PCR primers. For deletions under 1,100 bp, primers were designed outside of the breakpoints to amplify across the deletion sequence. For deletions where the reference allele was too large to be amplified by PCR, a double PCR approach was used. For the first PCR, one primer was designed within the putative deletion sequence while the other primer was placed external to the deletion breakpoint. PCR amplification using these primers would yield a product from the reference allele. For the second PCR, both primers flanked the putative deletion. Only samples that contained the deletion, would yield a product for this second PCR.

For duplication variants, since most duplications occur in a head to tail orientation, PCR primers were designed to amplify a product in this orientation. A forward direction primer was designed at the 3’ end of the duplicated sequence and a reverse primer was designed at the 5’ end of the duplicated sequence. These primers would amplify a product across the boundary at the duplication site. All PCR primer sequences were submitted to the Blast-like alignment tool (BLAT)^78^ to check for uniqueness of the sequence. When available, samples from three individuals reported as heterozygous for the variant were used for sequence validation along with one control (or reference) sample. When possible, samples from multiple families were used for validation.

Genomic DNA (∼50ng) was amplified using a SimpliAmp Thermal Cycler (Applied Biosystems) in a 20ul reaction volume with HotStarTaq Master Mix (Qiagen) in the presence of 2uM primers (IDT). The PCR conditions used were: 95°C 15min followed by 30 cycles of 95°C 20sec, 55°C 30sec, 72°C 2min with a final extension of 72°C 7min. The amplified PCR products were prepared for Sanger sequencing by adding ExoSAP-IT (USB) and incubating at 37°C for 45min followed by 80°C for 15min. The PCR products were then Sanger sequenced using the BigDye® Terminator v3.1 Cycle Sequencing kit (Part No. 4336917 Applied Biosystems). The sequencing reaction contained BigDye® Terminator v3.1 Ready Reaction Mix, 5X Sequencing Buffer, 5M Betaine solution (Part No. B0300 Sigma) and 0.64uM sequencing primer (IDT) in a total volume of 5ul. The sequencing reaction was performed in a SimpliAmp Thermal Cycler (Applied Biosystems) using the following program: 96°C 1min followed by 25 cycles of 96°C 10sec, 50°C 5sec, 60°C 1min15sec. The products were cleaned using XTerminator and SAM Solution (Applied Biosystems) with 30min of shaking at 1800rpm followed by centrifugation at 1000 rpm for 2min. The sequencing products were analyzed on a SeqStudio Genetic Analyzer (Applied Biosystems) and the sequencing traces were analyzed using Sequencher 5.4 (Gene Code)

### SVs on AD risk loci and AD genes

We first searched for SVs that are in linkage disequilibrium (LD) with AD associated loci from three GWASs^8–10^. There are 123 unique variants that reached genome-wide significance from three studies. After excluding nine variants that were not found in the WGS data, we searched for SVs that are in LD (R^2^ > 0.2) with the rest of 114 variants. For SVs, *P* value from fastGWA^42^ adjusting for PCs 1-5, age, sex, sequencing centers, sequencing platforms, and PCR status were also provided.

Then, we investigated SVs on known AD genes. A list of 20 expert curated AD risk/protective causal genes were downloaded from: https://adsp.niagads.org/index.php/gvc-top-hits-list. These genes were identified by a review of literature, pathway analysis, and by integration of genetic studies with myeloid genomics. All deletions, duplications, and inversions with missing rate less than 0.5 that overlap with these genes were inspected. Association of ultra-rare SVs on 20 AD genes were evaluated using SKAT-O test from R package SKAT^48^.

### Overall SV burden in AD

Overall SV burden between AD cases and controls was compared. SV burden was measured by the difference in the number of high-quality SVs in cases and controls. Logistic regression model adjusted for covariates (PCs 1-5, age, sex, sequencing center, sequencing platform, PCR status) were used. One-sided empirical p values (assuming increased SV burden in cases) were calculated based on 10,000 permutations. Particularly, we evaluated the burden of singletons and homozygous SVs in AD compared to controls.

### Association and functional analyses

In total, 136,092 SVs with a missing rate < 0.5 and minor allele count > 5 were evaluated using mixed linear model based tool (fastGWA) implemented in GCTA^42^. Age, sex, sample PCR status, sequencing platforms, sequencing centers, and PCs 1-5 calculated from common SNVs were included as covariates. The age of cases was determined by the age at disease onset. The age of controls was determined by the age at the last exam. Sparse genetic relationship matrix was generated using SNVs as well with a cutoff of 0.05. High-confident deletions, duplications, and inversions were selected by Samplot and experimentally validated by PCR. For insertions, only high-confident ones that are high-quality and not on the problematic regions were reported. Enrichment analysis for nominal significant signals (2,411 high-quality SVs with *P* < 0.05) was performed using clusterProfiler^79^.

Other than binary AD diagnosis, we also assessed SV association with cognitive scores, fluid biomarkers, and neuropathological measurements that were harmonized by the ADSP Phenotype Harmonization Consortium^80^. Cognitive scores include memory score (N = 6,413), executive function score (N = 5,762), language score (N = 6,130), and visuospatial score (N = 1,126)^80^. Fluid biomarkers include CSF Amyloid beta (N = 1,110), tau (N = 1,086), and P-tau (N = 1,087). Neuropathological measurements include Thal amyloid phases (N = 543), CERAD amyloid scores (N = 2,361), amyloid presence (dichotomous, N = 2,361), BRAAK tau phases (N = 2,357), ADNC severity scores (N = 540), NIA-REAGAN criteria for AD (N = 1,060).

## Declarations

## Ethics approval and consent to participate

## Consent for publication

Not applicable.

## Availability of data and materials

https://github.com/whtop/SV-ADSP-Pipeline https://dss.niagads.org/

## Competing interests

The authors declare that they have no competing interests.

## Funding

HW and PLC report grant support from RF1-AG074328 and P30-AG072979. AT, YQS, and JYT report grant support from RF1-AG074328. YYL reports grant support from U54-AG052427 and U24-AG041689. LSW reports grant support from U24-AG041689, U54-AG052427, U01-AG032984, U01-AG058654, and P30AG072979. LAF reports grant support from U54-AG052427, U01-AG058654, U01-AG062602, R01-AG048927, and P30-AG072978. WPL reports grant support from RF1-AG074328, P30-AG072979, U54-AG052427, and U24-AG041689.

## Data Availability

https://www.niagads.org/home

## Acknowledgements

The ADGC cohorts

The ADGC cohorts include Adult Changes in Thought (ACT) (U01 AG006781, U19 AG066567), the Alzheimer’s Disease Research Centers (ADRC) (P30 AG062429, P30 AG066468, P30 AG062421, P30 AG066509, P30 AG066514, P30 AG066530, P30 AG066507, P30 AG066444, P30 AG066518, P30 AG066512, P30 AG066462, P30 AG072979, P30 AG072972, P30 AG072976, P30 AG072975, P30 AG072978, P30 AG072977, P30 AG066519, P30 AG062677, P30 AG079280, P30 AG062422, P30 AG066511, P30 AG072946, P30 AG062715, P30 AG072973, P30 AG066506, P30 AG066508, P30 AG066515, P30 AG072947, P30 AG072931, P30 AG066546, P20 AG068024, P20 AG068053, P20 AG068077, P20 AG068082, P30 AG072958, P30 AG072959), the Chicago Health and Aging Project (CHAP) (R01 AG11101, RC4 AG039085, K23 AG030944), Indiana Memory and Aging Study (IMAS) (R01 AG019771), Indianapolis Ibadan (R01 AG009956, P30 AG010133), the Memory and Aging Project (MAP) (R01 AG17917), Mayo Clinic (MAYO) (R01 AG032990, U01 AG046139, R01 NS080820, RF1 AG051504, P50 AG016574), Mayo Parkinson’s Disease controls (NS039764, NS071674, 5RC2HG005605), University of Miami (R01 AG027944, R01 AG028786, R01 AG019085, IIRG09133827, A2011048), the Multi-Institutional Research in Alzheimer’s Genetic Epidemiology Study (MIRAGE) (R01 AG09029, R01 AG025259), the National Centralized Repository for Alzheimer’s Disease and Related Dementias (NCRAD) (U24 AG021886), the National Institute on Aging Late Onset Alzheimer’s Disease Family Study (NIA-LOAD) (U24 AG056270), the Religious Orders Study (ROS) (P30 AG10161, R01 AG15819), the Texas Alzheimer’s Research and Care Consortium (TARCC) (funded by the Darrell K Royal Texas Alzheimer’s Initiative), Vanderbilt University/Case Western Reserve University (VAN/CWRU) (R01 AG019757, R01 AG021547, R01 AG027944, R01 AG028786, P01 NS026630, and Alzheimer’s Association), the Washington Heights-Inwood Columbia Aging Project (WHICAP) (RF1 AG054023), the University of Washington Families (VA Research Merit Grant, NIA: P50AG005136, R01AG041797, NINDS: R01NS069719), the Columbia University Hispanic Estudio Familiar de Influencia Genetica de Alzheimer (EFIGA) (RF1 AG015473), the University of Toronto (UT) (funded by Wellcome Trust, Medical Research Council, Canadian Institutes of Health Research), and Genetic Differences (GD) (R01 AG007584). The CHARGE cohorts are supported in part by National Heart, Lung, and Blood Institute (NHLBI) infrastructure grant HL105756 (Psaty), RC2HL102419 (Boerwinkle) and the neurology working group is supported by the National Institute on Aging (NIA) R01 grant AG033193.

The CHARGE cohorts

The CHARGE cohorts participating in the ADSP include the following: Austrian Stroke Prevention Study (ASPS), ASPS-Family study, and the Prospective Dementia Registry-Austria (ASPS/PRODEM-Aus), the Atherosclerosis Risk in Communities (ARIC) Study, the Cardiovascular Health Study (CHS), the Erasmus Rucphen Family Study (ERF), the Framingham Heart Study (FHS), and the Rotterdam Study (RS). ASPS is funded by the Austrian Science Fond (FWF) grant number P20545-P05 and P13180 and the Medical University of Graz. The ASPS-Fam is funded by the Austrian Science Fund (FWF) project I904), the EU Joint Programme – Neurodegenerative Disease Research (JPND) in frame of the BRIDGET project (Austria, Ministry of Science) and the Medical University of Graz and the Steiermärkische Krankenanstalten Gesellschaft. PRODEM-Austria is supported by the Austrian Research Promotion agency (FFG) (Project No. 827462) and by the Austrian National Bank (Anniversary Fund, project 15435. ARIC research is carried out as a collaborative study supported by NHLBI contracts (HHSN268201100005C, HHSN268201100006C, HHSN268201100007C, HHSN268201100008C, HHSN268201100009C, HHSN268201100010C, HHSN268201100011C, and HHSN268201100012C). Neurocognitive data in ARIC is collected by U01 2U01HL096812, 2U01HL096814, 2U01HL096899, 2U01HL096902, 2U01HL096917 from the NIH (NHLBI, NINDS, NIA and NIDCD), and with previous brain MRI examinations funded by R01-HL70825 from the NHLBI. CHS research was supported by contracts HHSN268201200036C, HHSN268200800007C, N01HC55222, N01HC85079, N01HC85080, N01HC85081, N01HC85082, N01HC85083, N01HC85086, and grants U01HL080295 and U01HL130114 from the NHLBI with additional contribution from the National Institute of Neurological Disorders and Stroke (NINDS). Additional support was provided by R01AG023629, R01AG15928, and R01AG20098 from the NIA. FHS research is supported by NHLBI contracts N01-HC-25195 and HHSN268201500001I. This study was also supported by additional grants from the NIA (R01s AG054076, AG049607 and AG033040 and NINDS (R01 NS017950). The ERF study as a part of EUROSPAN (European Special Populations Research Network) was supported by European Commission FP6 STRP grant number 018947 (LSHG-CT-2006-01947) and also received funding from the European Community’s Seventh Framework Programme (FP7/2007-2013)/grant agreement HEALTH-F4-2007-201413 by the European Commission under the programme “Quality of Life and Management of the Living Resources” of 5th Framework Programme (no. QLG2-CT-2002-01254). High-throughput analysis of the ERF data was supported by a joint grant from the Netherlands Organization for Scientific Research and the Russian Foundation for Basic Research (NWO-RFBR 047.017.043). The Rotterdam Study is funded by Erasmus Medical Center and Erasmus University, Rotterdam, the Netherlands Organization for Health Research and Development (ZonMw), the Research Institute for Diseases in the Elderly (RIDE), the Ministry of Education, Culture and Science, the Ministry for Health, Welfare and Sports, the European Commission (DG XII), and the municipality of Rotterdam. Genetic data sets are also supported by the Netherlands Organization of Scientific Research NWO Investments (175.010.2005.011, 911-03-012), the Genetic Laboratory of the Department of Internal Medicine, Erasmus MC, the Research Institute for Diseases in the Elderly (014-93-015; RIDE2), and the Netherlands Genomics Initiative (NGI)/Netherlands Organization for Scientific Research (NWO) Netherlands Consortium for Healthy Aging (NCHA), project 050-060-810. All studies are grateful to their participants, faculty and staff. The content of these manuscripts is solely the responsibility of the authors and does not necessarily represent the official views of the National Institutes of Health or the U.S. Department of Health and Human Services.

## Authors’ contribution

HW, YYL, JF, JSM, LSW, BNV, LAF, GDS, and WPL performed variant detection, quality check, and genotype/phenotype acquisition. HW, AT, YQS, JYT, and WPL performed statistical analyses. BAD, PLC, and GDS performed experimental validation. HW, BAD, PLC, AT, YQS, JF, JYT, YYL, JSM, BNV, LAF, GDS, and WPL interpreted results. HW and WPL wrote the first draft of the manuscript. All authors read, critically revised, and approved the manuscript.

## Notes

### Competing Interest Statement

The authors have declared no competing interest.

