## Supplementary Figures for "Structural Variation Detection and Association Analysis of Whole-Genome-Sequence Data from 16,905 Alzheimer’s Diseases Sequencing Project Subjects"

^1^Department of Pathology and Laboratory Medicine, Perelman School of Medicine, University of Pennsylvania, PA 19104, USA, ^2^Penn Neurodegeneration Genomics Center, Perelman School of Medicine, University of Pennsylvania, PA 19104, USA, ^3^Bioinformatics Research Center, North Carolina State University, NC 27695, USA, ^4^Department of Medicine (Biomedical Genetics), Boston University School of Medicine, MA 02118, USA, ^5^Department of Surgery, Scholl of Medicine, University of Colorado, CO 80045, USA, ^6^Taub Institute for Research on Alzheimer’s Disease and the Aging Brain, College of Physicians and Surgeons, Columbia University, NY 10032, USA, ^7^Department of Neurology, College of Physicians and Surgeons, Columbia University and the New York Presbyterian Hospital, NY 10032, USA, ^8^Department of Neurology, Boston University School of Medicine, MA 02118, USA, ^9^Department of Ophthalmology, Boston University School of Medicine, MA 02118, USA, ^10^Department of Biostatistics, Boston University School of Public Health, MA 02118, USA, ^11^Department of Epidemiology, Boston University School of Public Health, MA 02118, USA

**Search Terms:** **Alzheimer's disease, Structural variation, Copy number variation**

Supplementary Figures

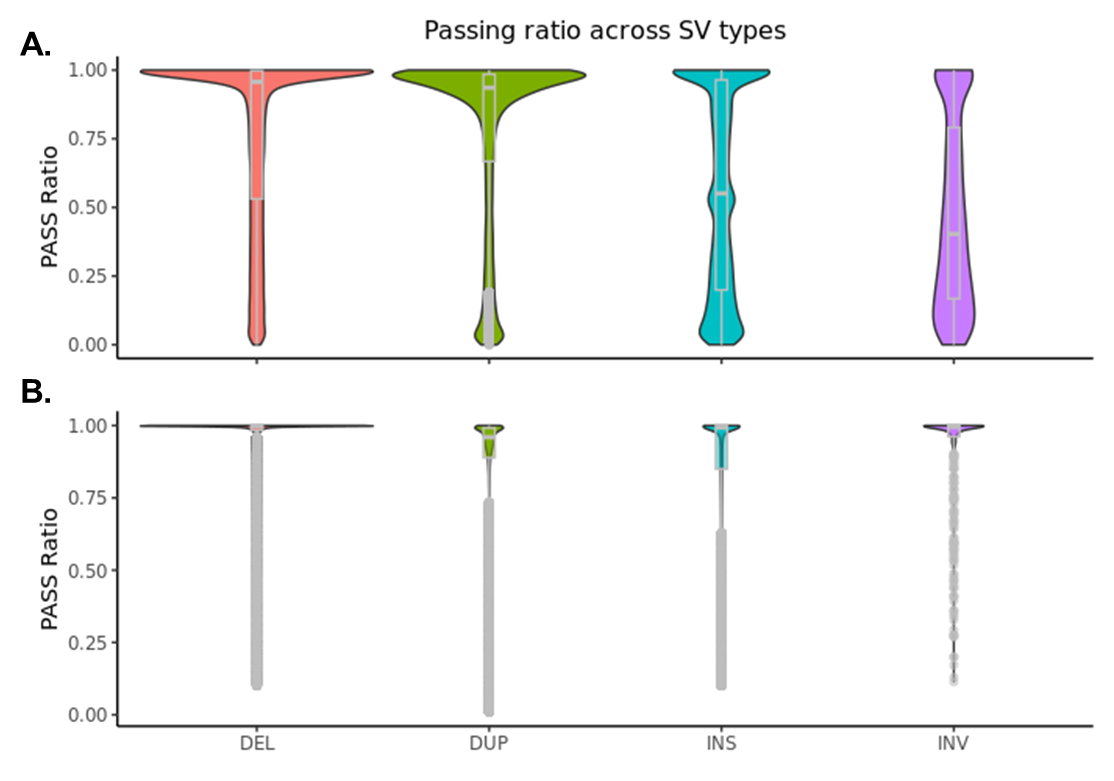

Figure S1. The ratio of passed calls for each type of SVs

A. For all 400,234 SVs. B. For 168,223 high-quality SVs.

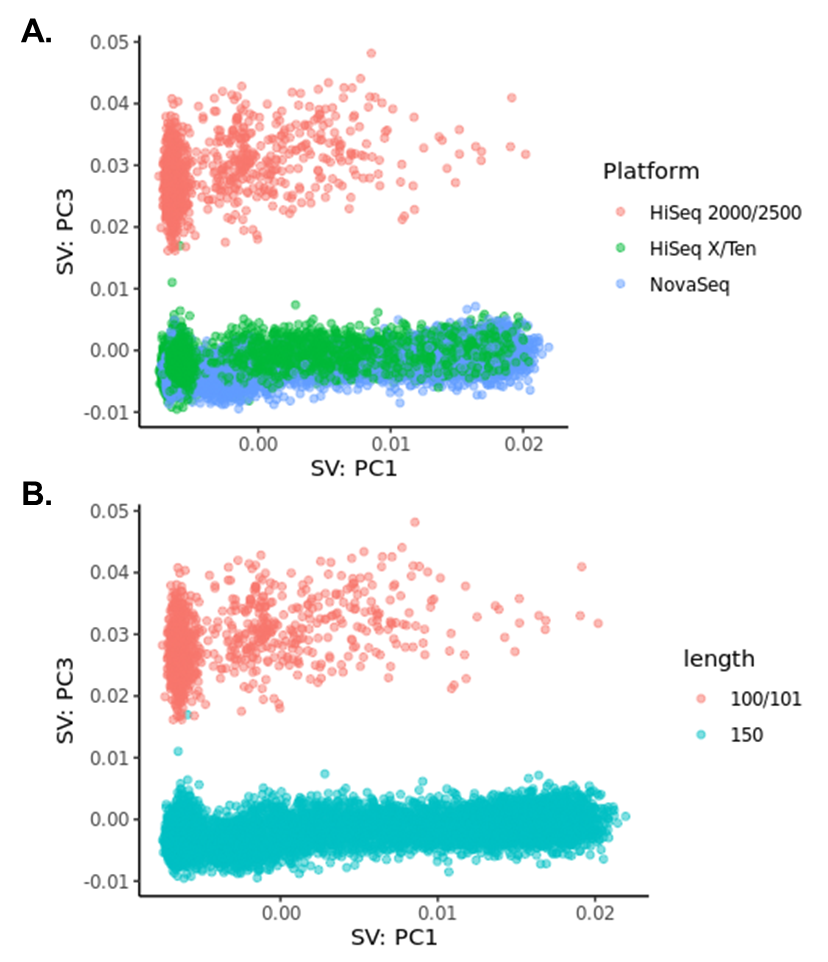

Figure S2. Principal component analysis (PCA) of SVs

**A.** PC1 and PC3 colored by sequencing platform **B.** PC1 and PC3 colored by read length

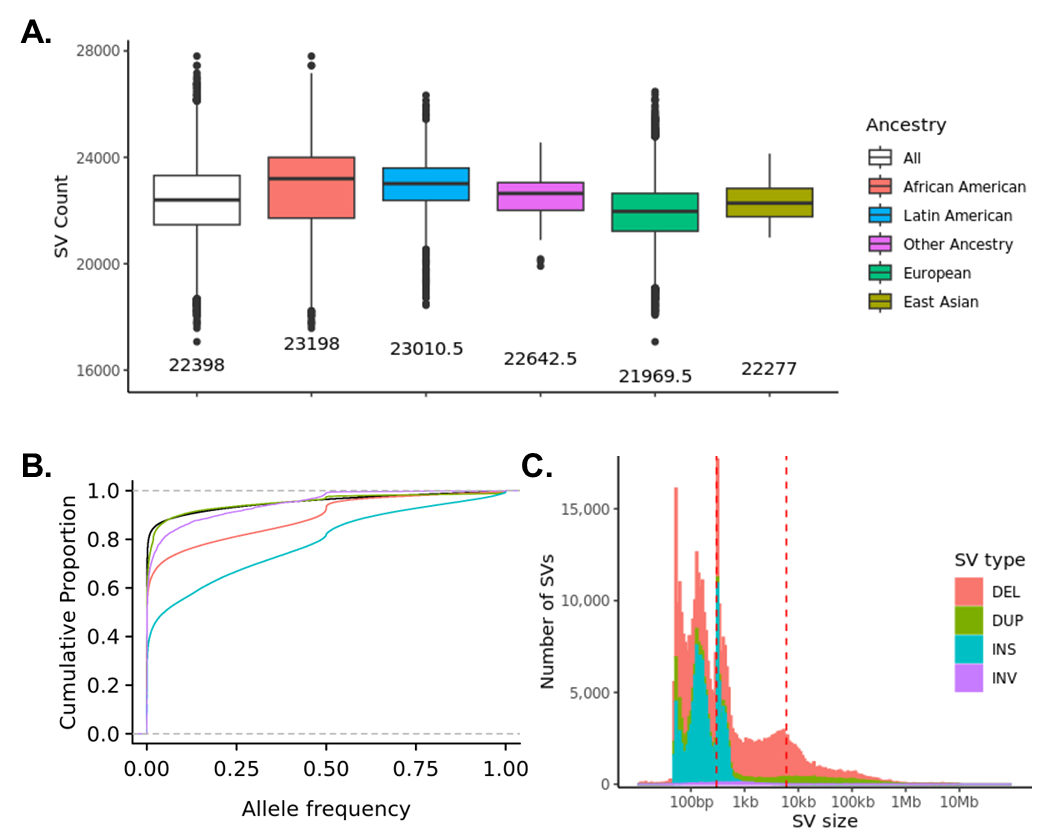

Figure S3. SV characteristics

**A.** Number of SVs per individual by ancestry. **B.** Allele frequency distribution of all types of SVs compared to SNVs. **C.** Distribution of SV size for all SVs.

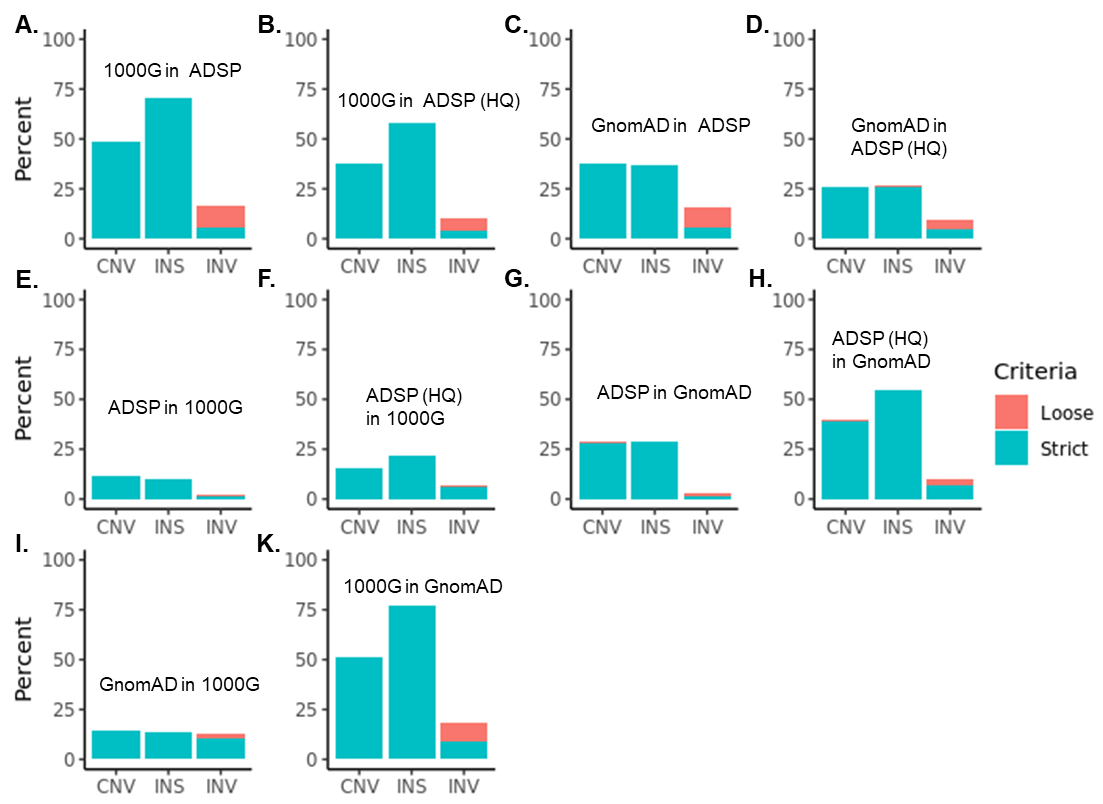

Figure S4. Comparison of SVs with GnomAD and 1000 Genome

**A.** Percent of SVs from 1000 Genome that are in ADSP. **B.** Percent of SVs from 1000 Genome that are in ADSP high-quality SVs. **C.** Percent of SVs from GnomAD that are in ADSP. **D.** Percent of SVs from GnomAD that are in ADSP high-quality SVs. **E.** Percent of SVs from ADSP that are in 1000 Genome. **F.** Percent of high-quality SVs from ADSP that are in 1000 Genome. **G.** Percent of SVs from ADSP that are in GnomAD. **H.** Percent of high-quality SVs from ADSP that are in GnomAD. **I.** Percent of SVs from GnomAD that are in 1000 Genome. **K.** Percent of SVs from 1000 Genome that are in GnomAD. With loose match, SVs are matched to all types of SVs in the same location. HQ, high-quality; CNV, copy number variant; INS, insertion; INV, inversion.

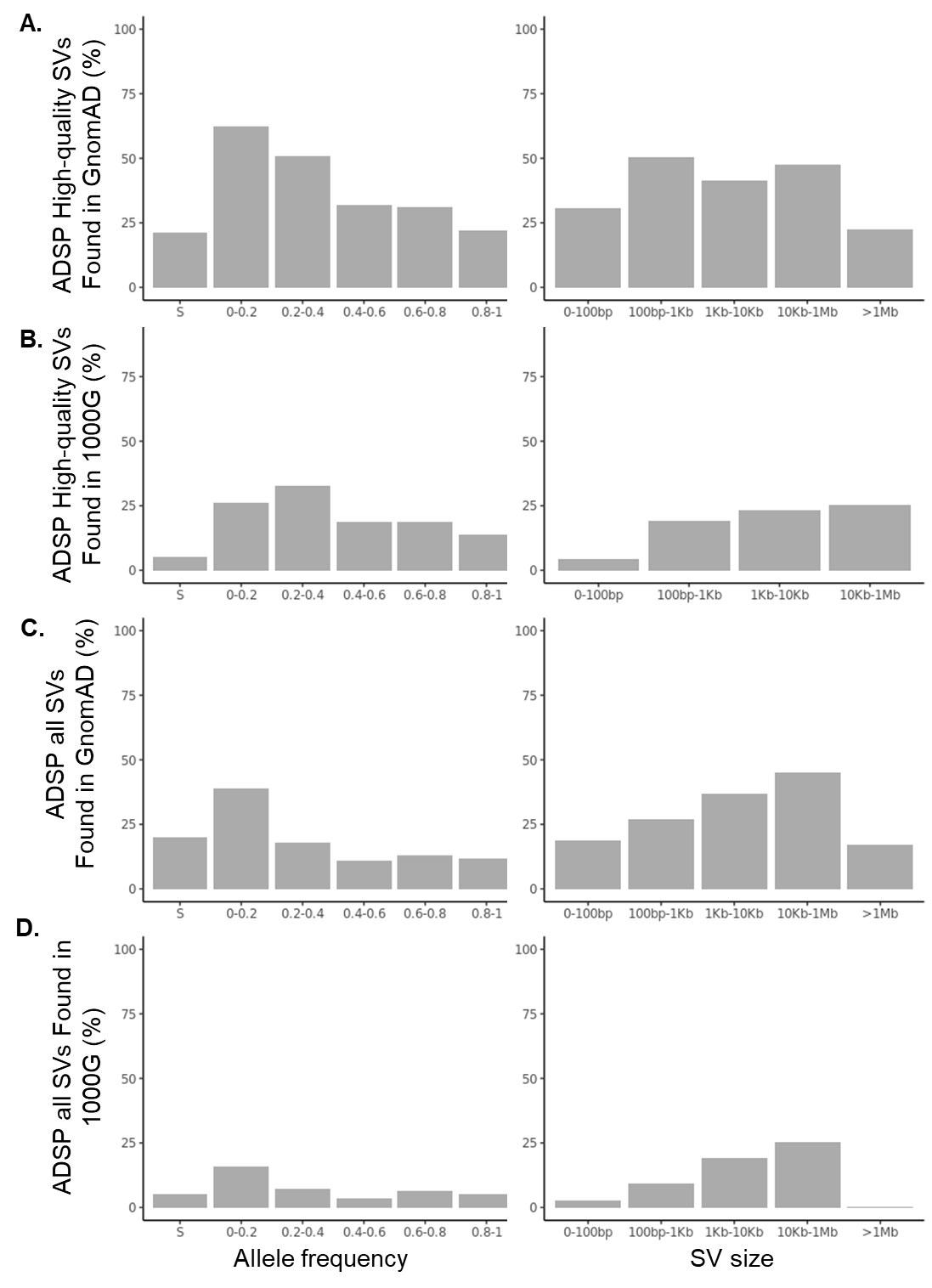

**Figure S5.** Comparison ADSP SVs with GnomAD and 1000 Genome

**A.** Percent of ADSP high-quality SVs that are in GnomAD by SV allele frequency and SV size (S represents singletons). **B.** Percent of ADSP high-quality SVs that are in 1000 Genome by SV allele frequency and SV size. **C.** Percent of ADSP all SVs that are in GnomAD by SV allele frequency and SV size. **D.** Percent of ADSP all SVs that are in 1000 Genome by SV allele frequency and SV size.

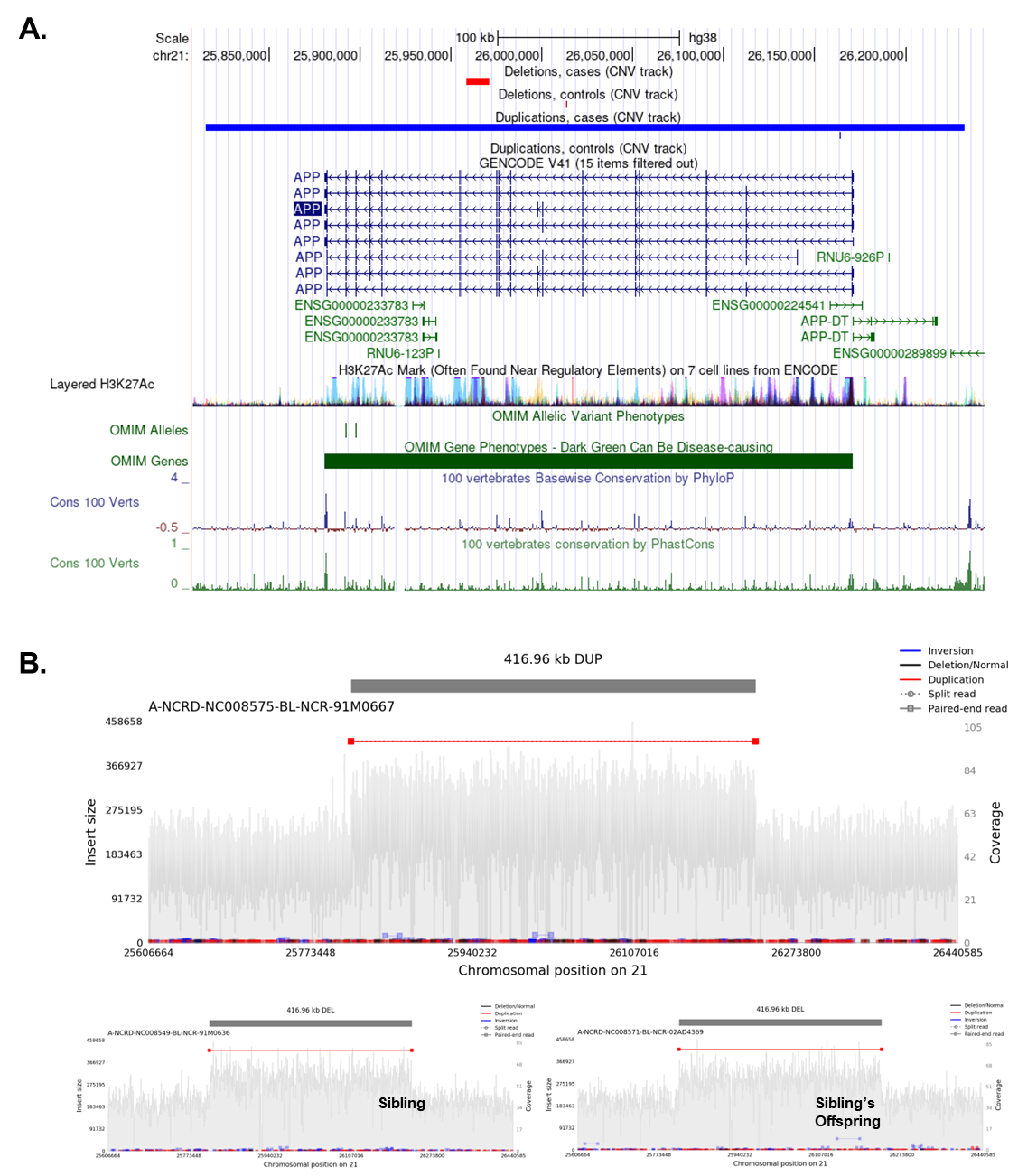

Figure S6. Ultra-rare SVs on *APP*

**A.** SVs with MAC < 5 on *APP*. **B.** Samplot for one individual with the 400kb duplication covering entire *APP*. The Samplot for the 400kb region were also plotted for his two sisters.

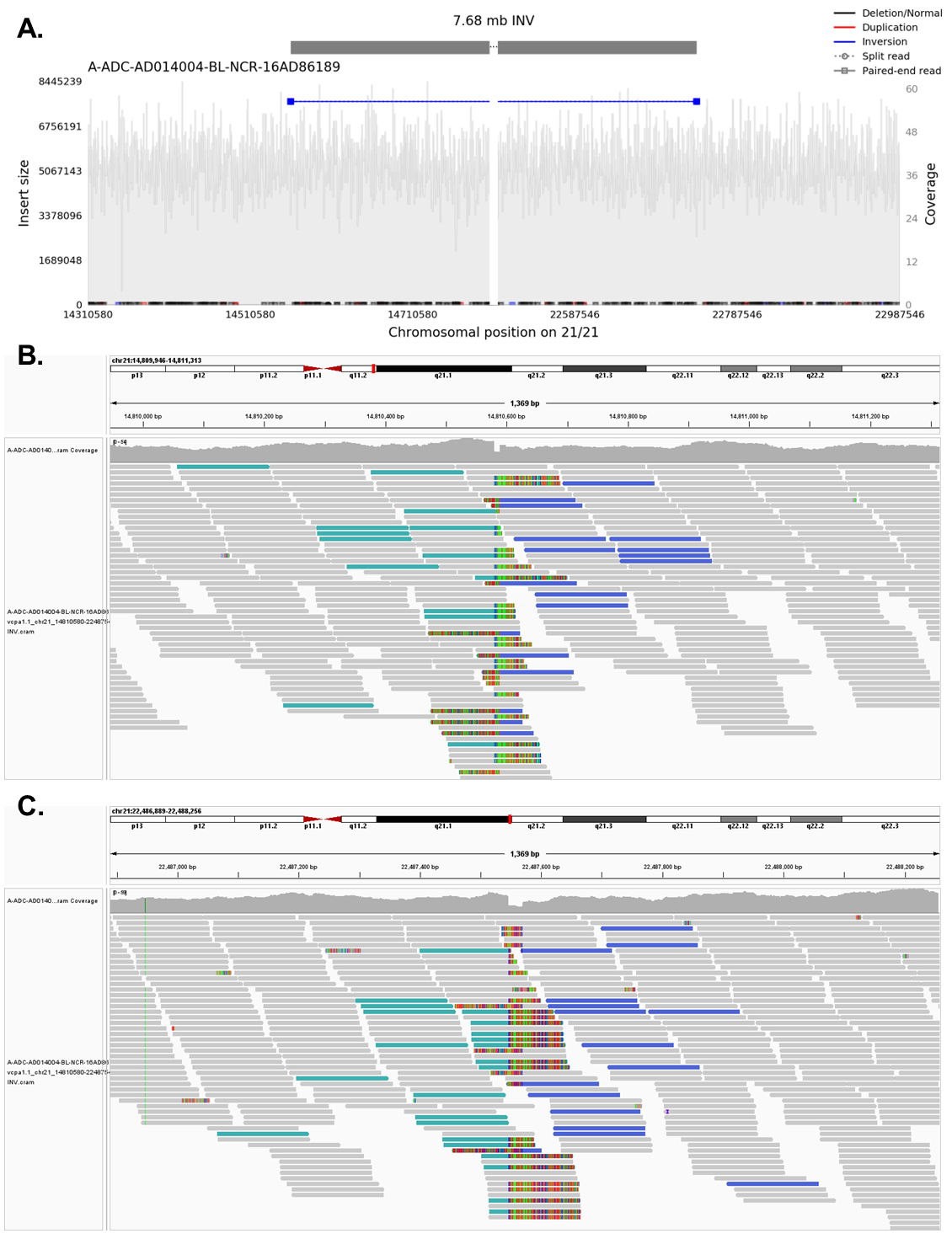

Figure S7. The 7.68 Mb inversion covering 21q21.2

**A.** Alignment view (using samplot) of the inversion. **B.** The left breakpoint of the inversion. Soft clips map to the right of the inversion. **C.** The right breakpoint of the inversion. Soft clips map to the left of the inversion. Green: paired reads are both on positive strand. Blue: paired reads are both on negative strand. Note that the inversion was confirmed by experimental validation.

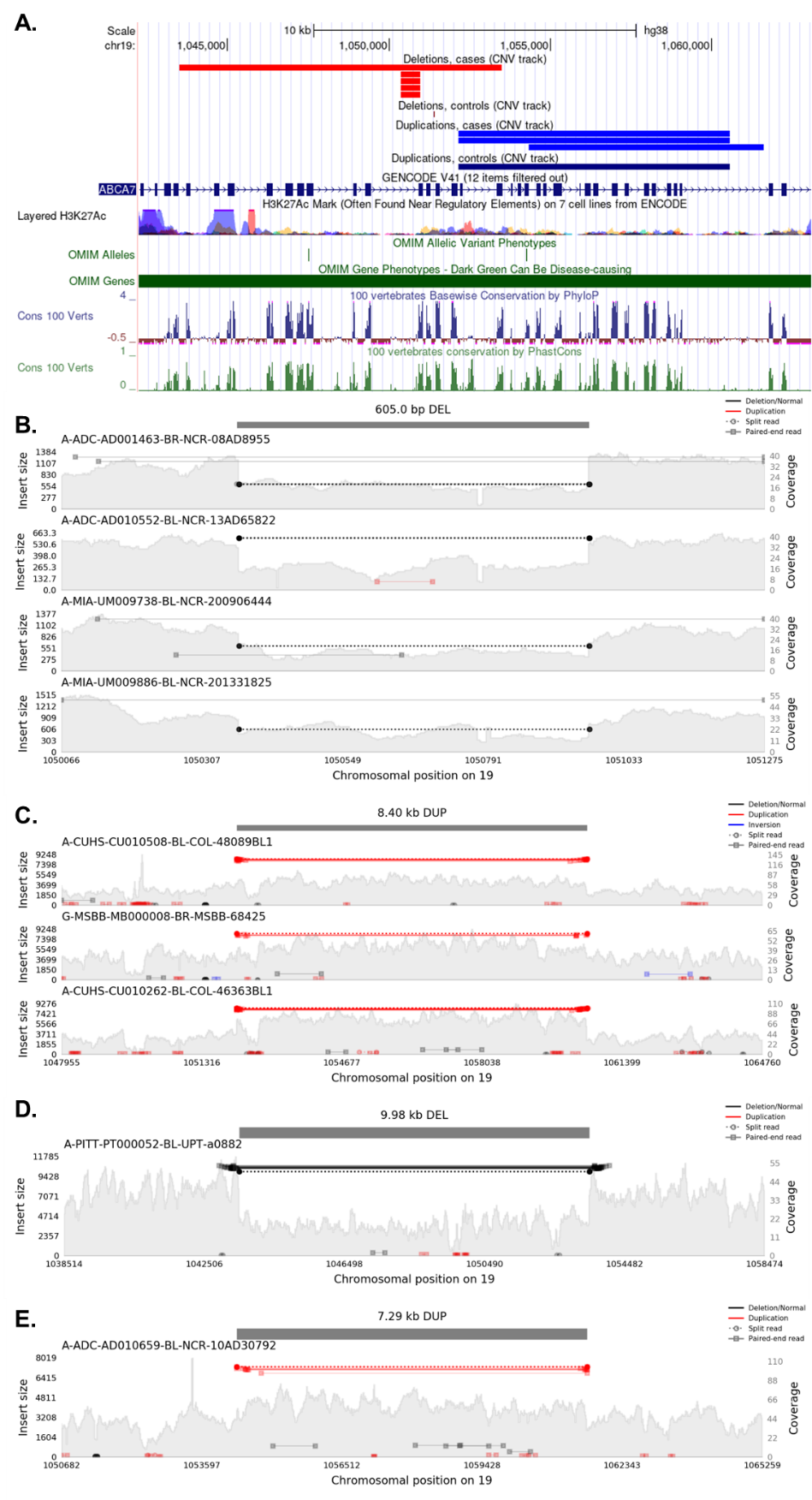

Figure S8. Ultra-rare SVs on *ABCA7*

**A.** SVs with MAC < 5 on *ABCA7*. **B-E.** Alignment view (using samplot) of non-intronic deletions and duplications on *ABCA7*.

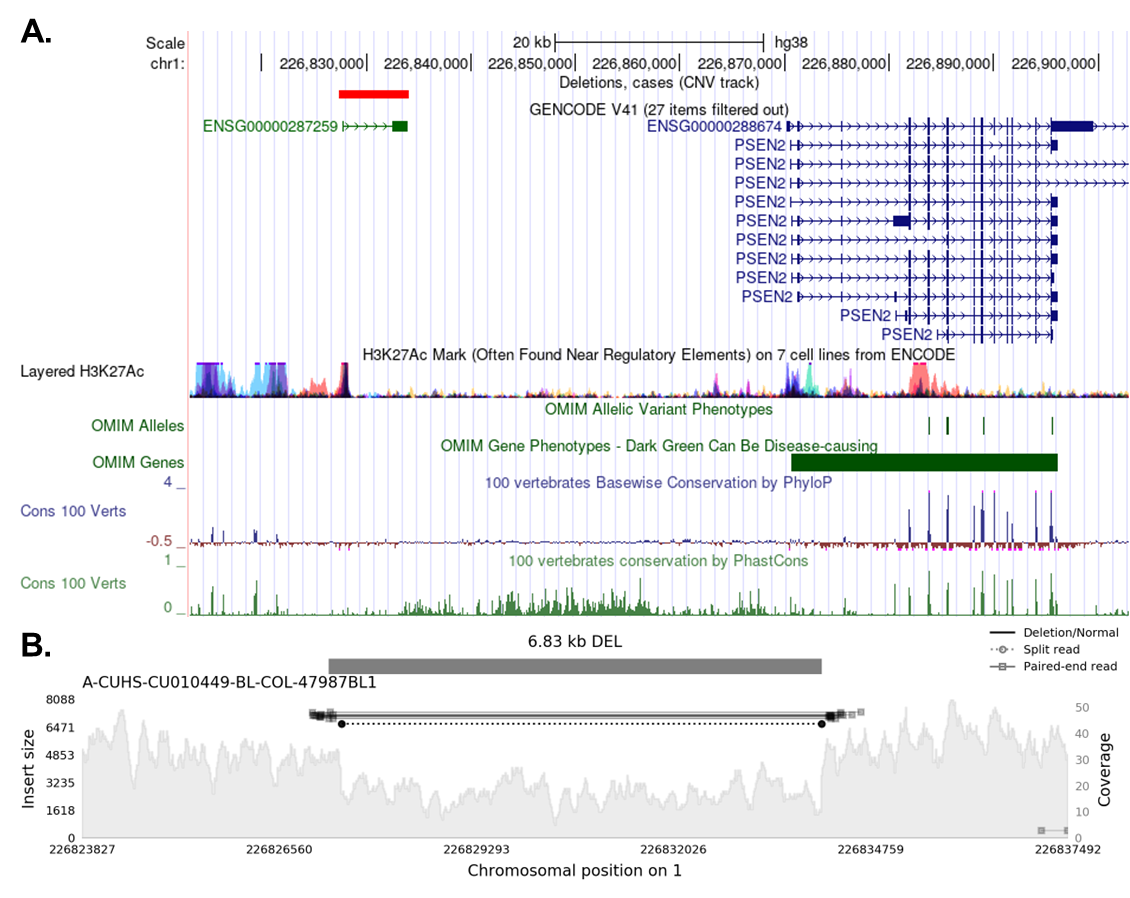

Figure S9. Singleton deletion on *lnc-PSEN2-7*

**A.** Singleton deletion of *lnc-PSEN2-7* on genome brower. **B.** Alignment view (using samplot) of singleton deletion on *lnc-PSEN2-7*.

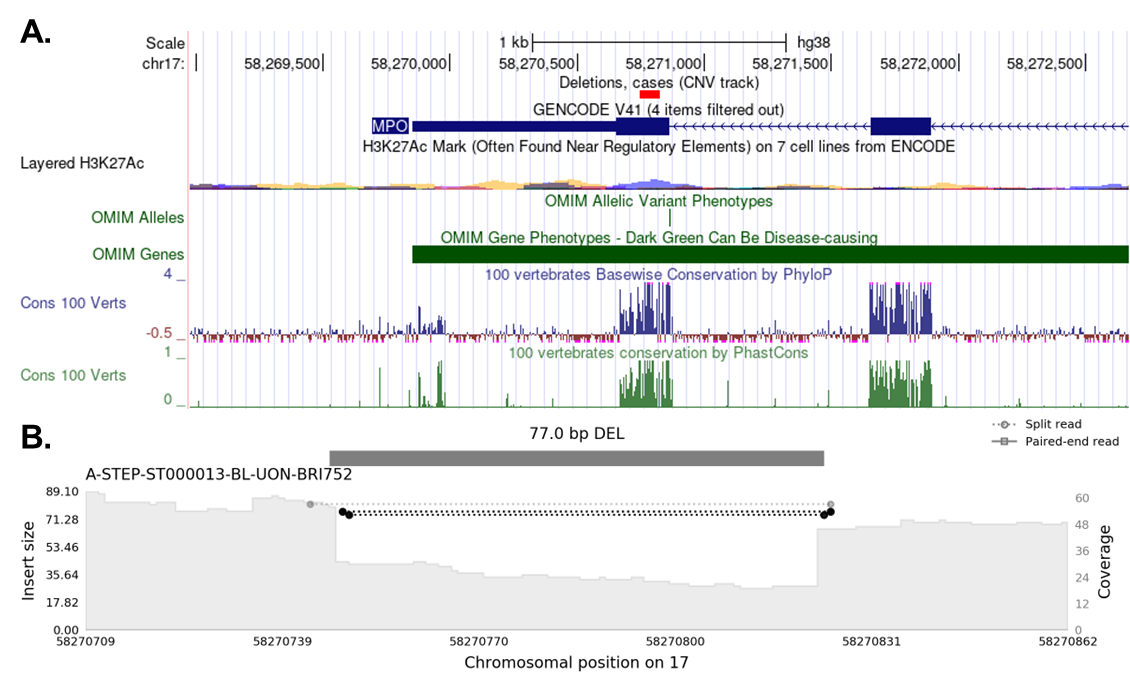

Figure S10. Singleton deletion on *MPO*

**A.** Singleton deletion of *MPO* on genome browser. **B.** Alignment view (using samplot) of singleton deletion on *MPO*.

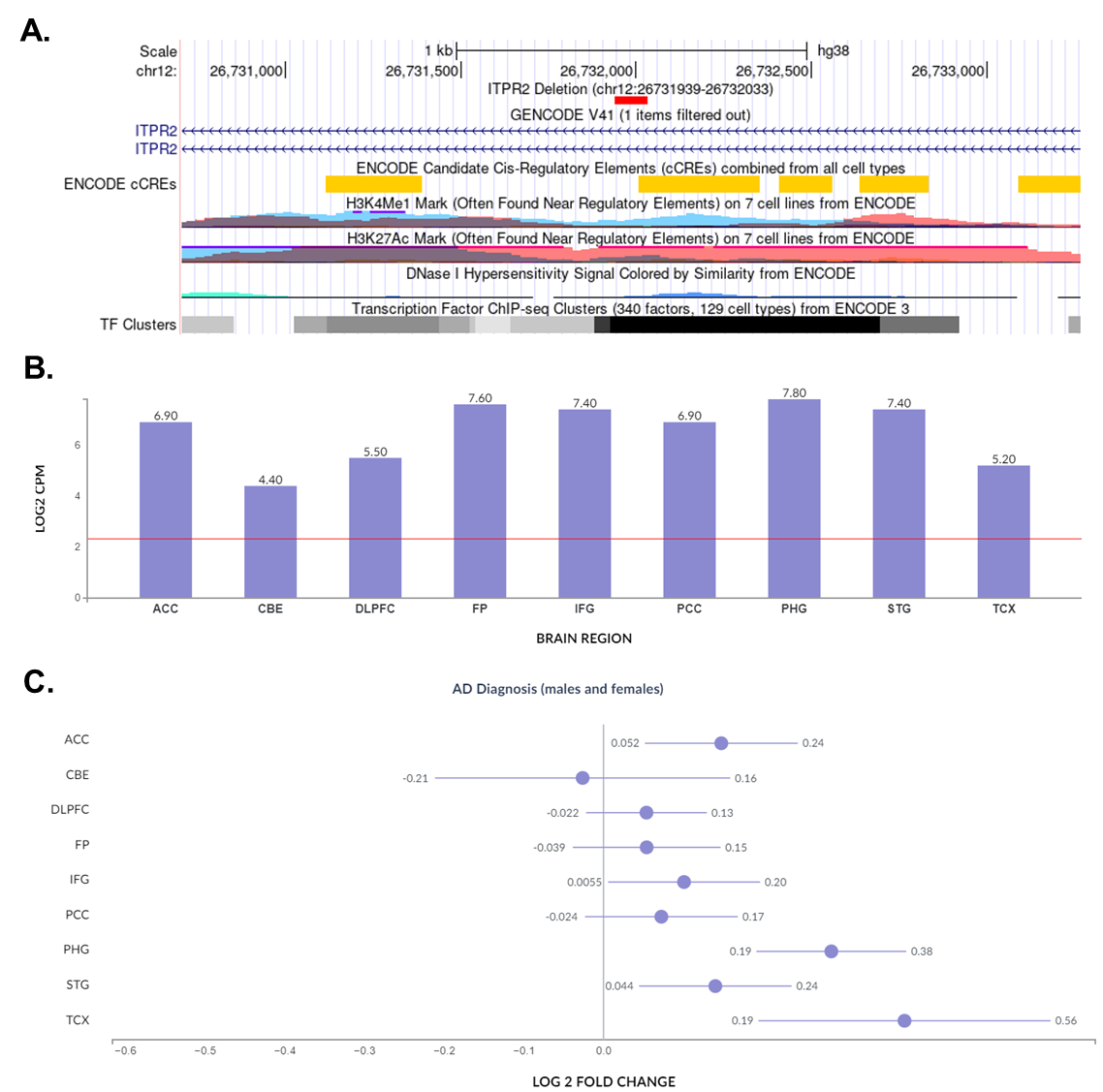

Figure S11. Genomic location of *ITPR2* intronic deletion and *ITPR2* expression

**A.** The deletion region (chr12:26731939-26732033) in genome browser. **B.** *ITPR2* expression across brain regions. **C.** Fold change of *ITPR2* expression in cases and controls.

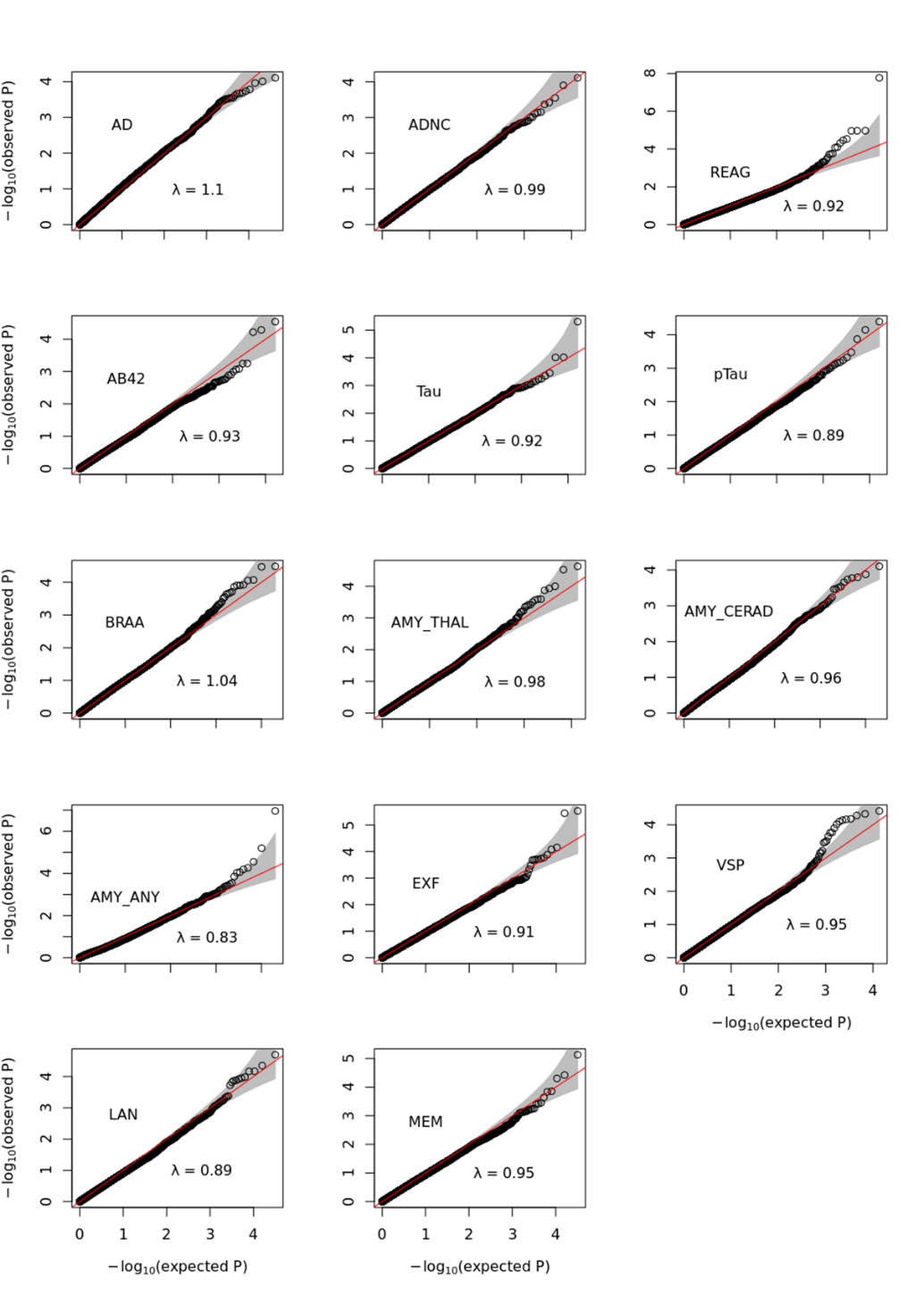

Figure S12. QQ plot of association analysis

AD, Alzheimer’s disease; ADNC, AD neuropathological change severity score; REAG, NIA-Reagan diagnosis of AD; AB42, cerebrospinal fluid (CSF) β-amyloid (1-42); Tau, CSF tau; pTau, CSF phosphorylated tau; BRAA, Braak staging; AMY_THAL, Thal amyloid phases; AMY_CERAD, CERAD amyloid score; AMY_ANY, amyloid presence (dichotomous); EXF, executive function score; VSP, visuospatial score; LAN, language score; MEM, memory score.

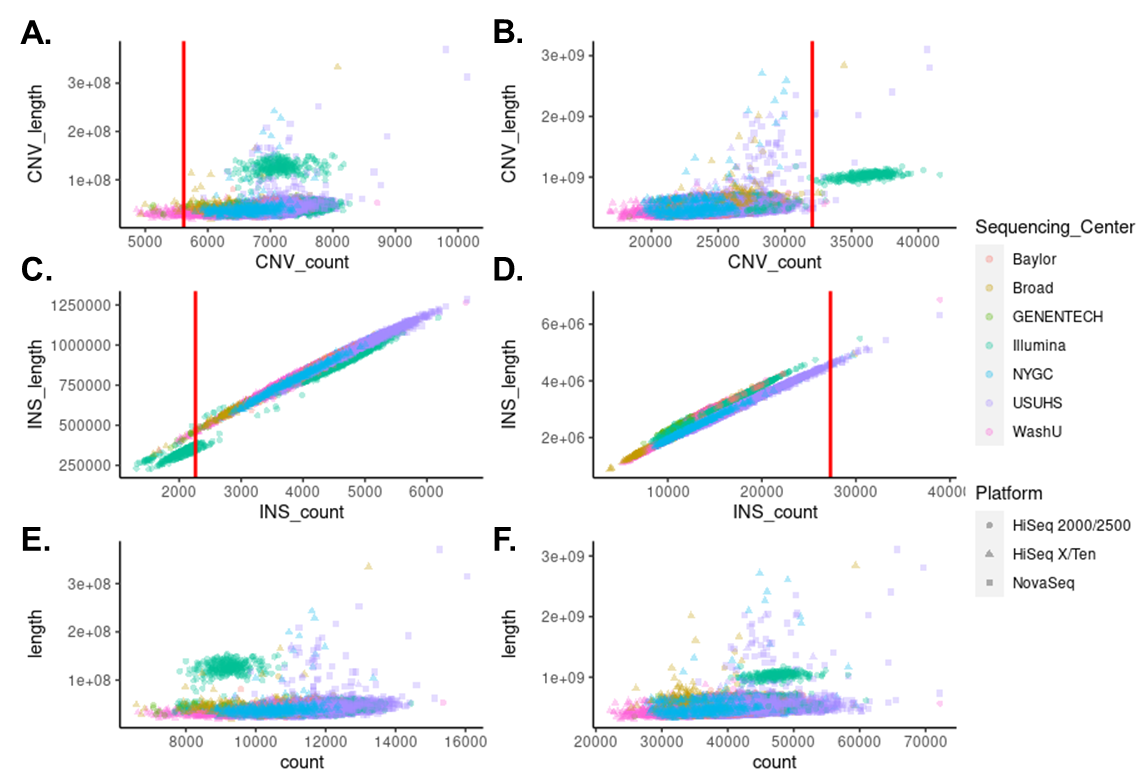

Figure S13. Scatter plot by SV count and SV length. Each dot represents one sample

**A.** High-quality copy number variants (CNVs, deletions and duplications). Red line represents median count – 4 × mean absolute deviation (MAD). **B.** All CNVs. Red line represents median count + 4 × mean absolute deviation (MAD). **C.** High-quality insertions. Red line represents median count – 4 × MAD. **D.** All insertions. Red line represents median count + 4 × MAD. **E.** High-quality SVs. **F.** All SVs.

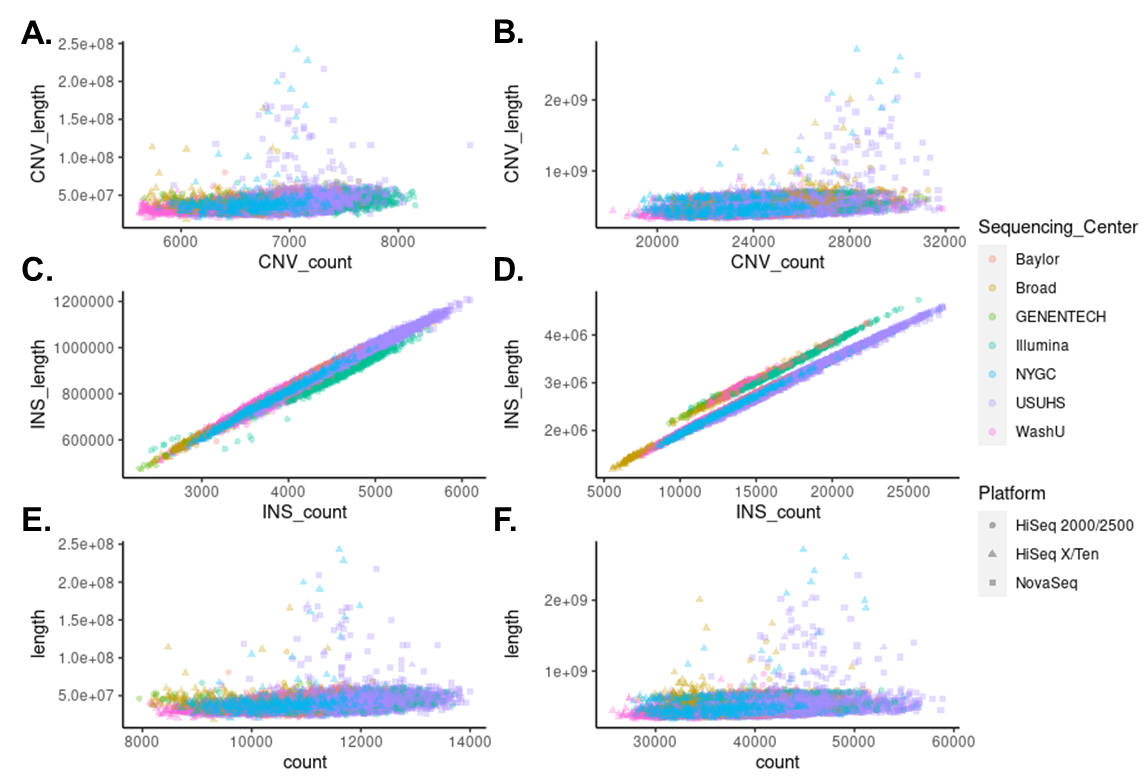

Figure S14. Scatter plot by SV count and SV length after removing outlier samples.

Each dot represents one sample. **A.** High-quality copy number variants (CNVs). **B.** All CNVs. **C.** High-quality insertions. **D.** All insertions. **E.** High-quality SVs. **F.** All SVs.

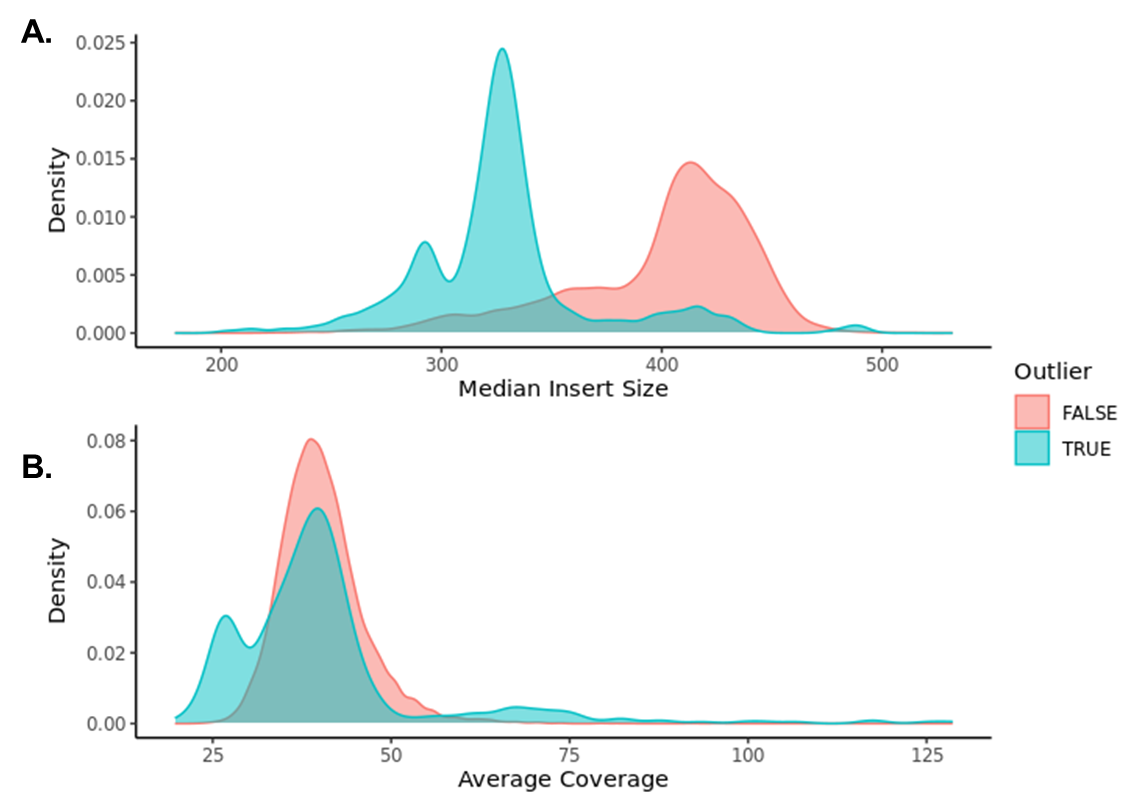

Figure S15. Distribution of median insert size and average coverage for 12908 samples and 463 outliers

**A.** Distribution of median insert size. **B.** Distribution of average coverage.

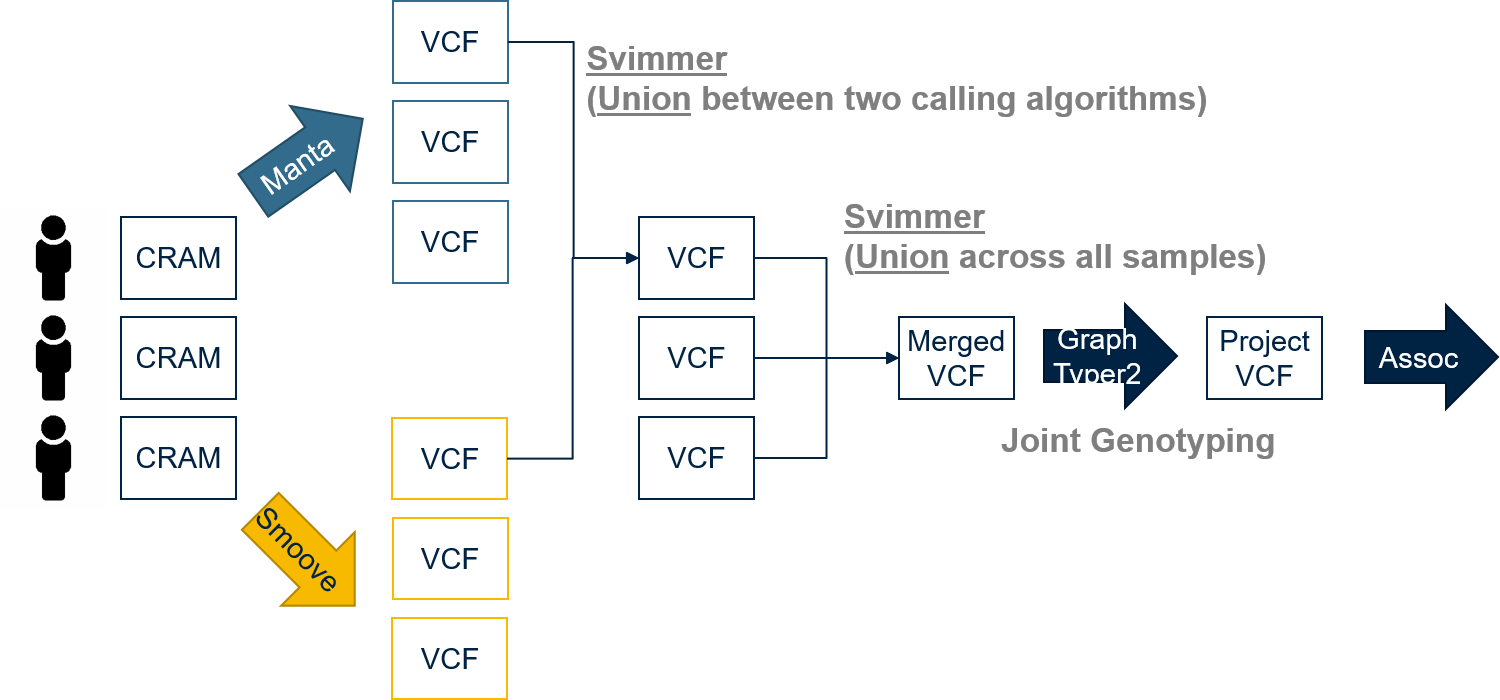

Figure S16. The pipeline of SV detection

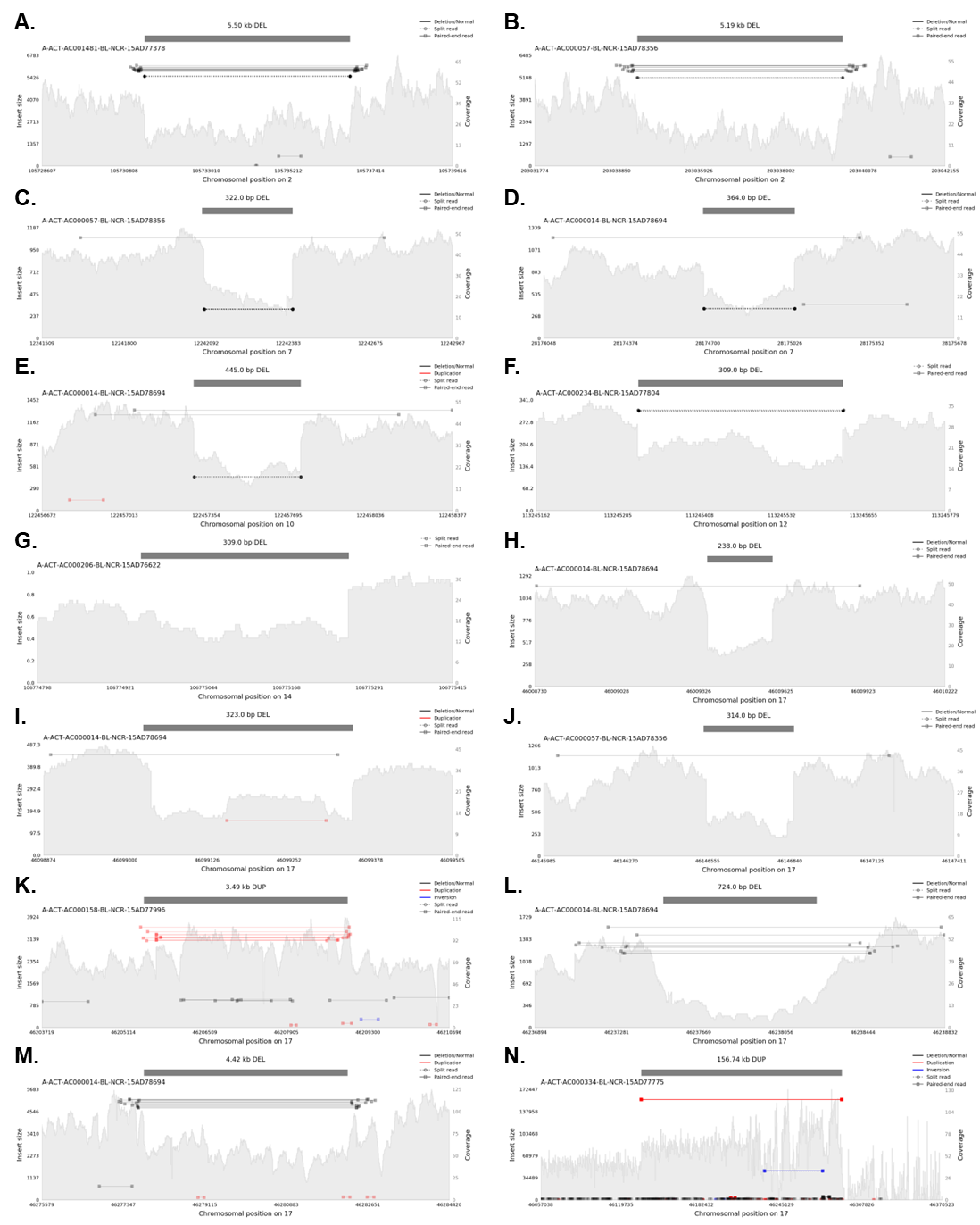

Figure S17. Samplot for deletions and duplications in Table 2

**A-N** represents Samplot for one selected sample for chr2:105731359-105736864:DEL, chr2:203034369-203039560:DEL, chr7:12242077-12242399:DEL, chr7:28174681-28175045:DEL, chr10:122457302-122457747:DEL, chr12:113245316-113245625:DEL, chr14:106774952-106775261:DEL, chr17:46009357-46009595:DEL, chr17:46099028-46099351:DEL, chr17:46146541-46146855:DEL, chr17:46205463-46208952:DUP, chr17:46237501-46238225:DEL, chr17:46277789-46282210:DEL, and chr17:46135409-46292152:DUP, respectively.

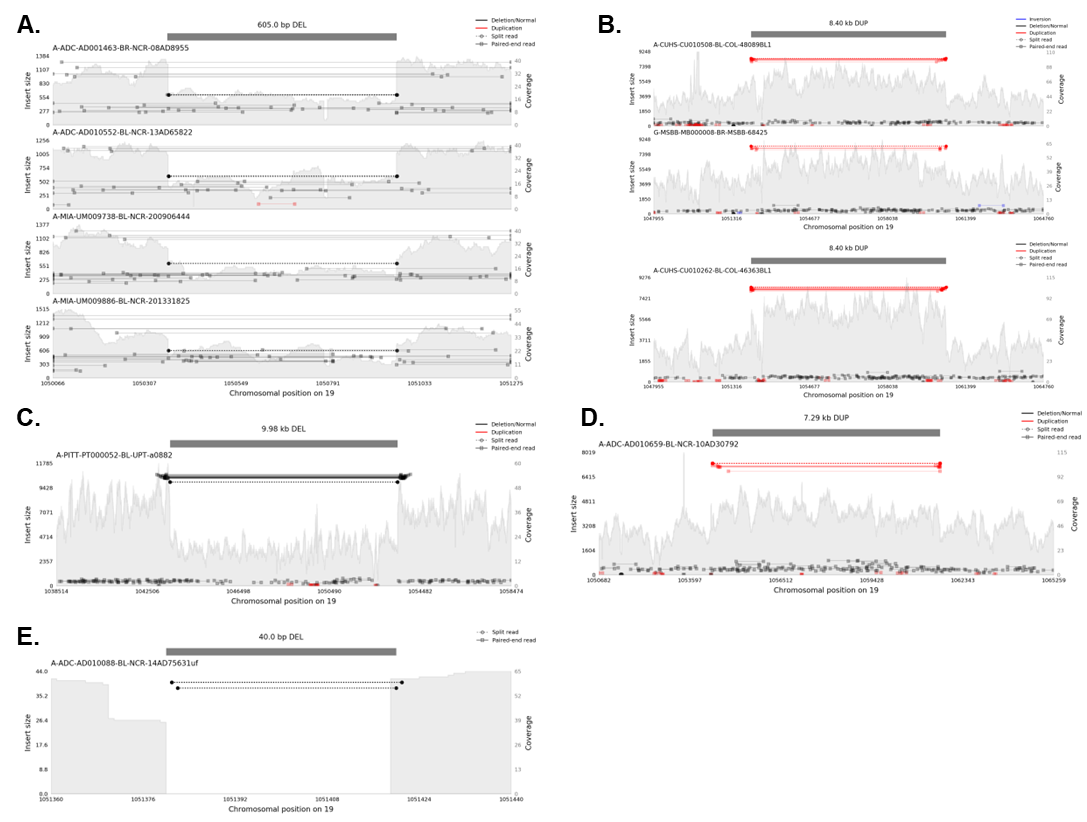

Figure S18. Ultra-rare CNVs on ABCA7

**A.** Deletion on chr19:1050368-1050973. **B.** Heterozygous duplication (top) and homozygous duplication (bottom) on chr19:1052156-1060559. **C-E.** Deletion on chr19:1043504-1053484, duplication on chr19:1054326-1061615, and deletion on chr19:1051380-1051420.

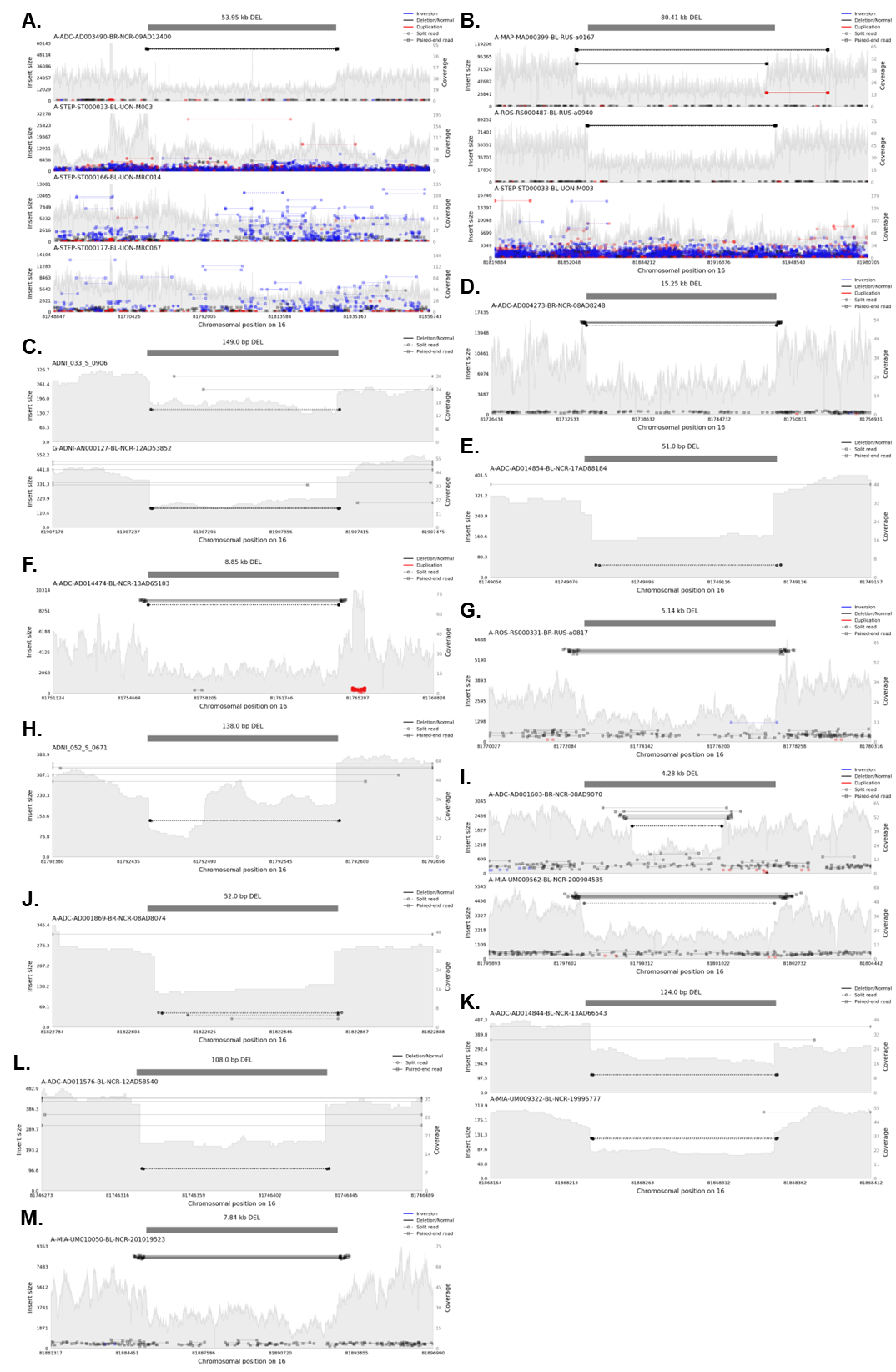

Figure S19. Ultra-rare CNVs on PLCG2

**A-M.** Deletion on chr16:81775821-81829769, deletion on chr16:81860089-81940500, deletion on chr16:81907252-81907401, deletion on chr16:81734058-81749307, deletion on chr16:81749081-81749132, deletion on chr16:81755550-81764402, deletion on chr16:81772599-81777744, deletion on chr16:81792449-81792587, deletion on chr16:81798030-81802305, deletion on chr16:81822810-81822862, deletion on chr16:81868226-81868350, deletion on chr16:81746327-81746435, and deletion on chr16:81885235-81893072.

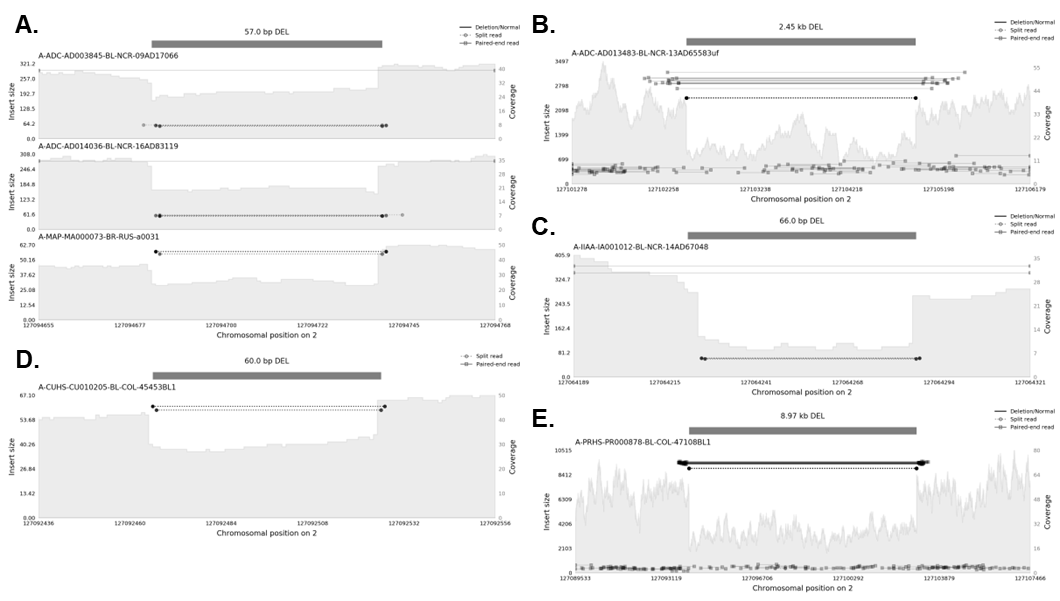

Figure S20. Ultra-rare CNVs on BIN1

**A-E.** Deletion on chr2:127094683-127094740, deletion on chr2:127102503-127104954, deletion on chr2:127064222-127064288, deletion on chr2:127092466-127092526, deletion on chr2:127094016-127102983.

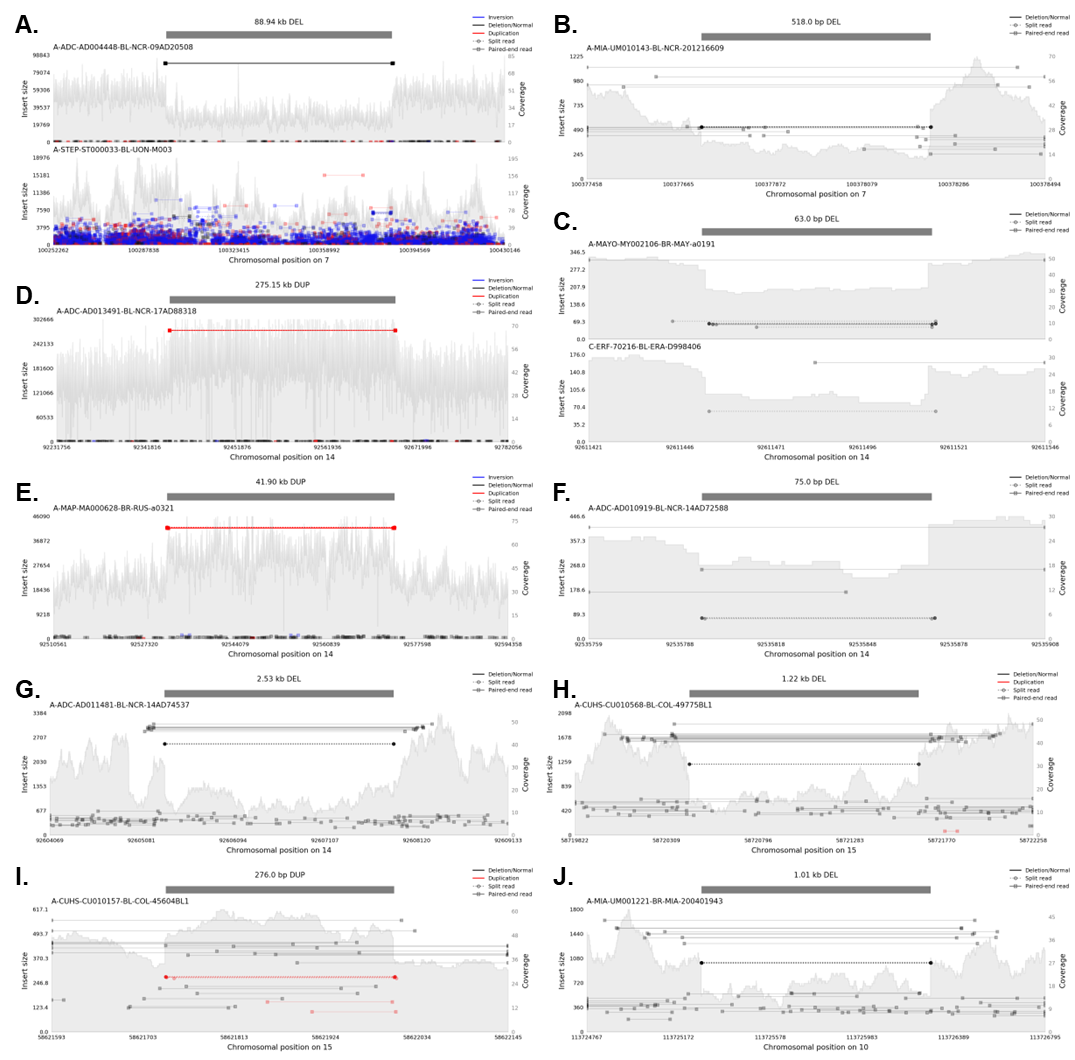

Figure S21. Ultra-rare CNVs on PILRA, RIN3, ADAM10 and CASP7

**A-B.** Deletions on *PILRA*: chr7:100296733-100385675, and chr7:100377717-100378235. **C-G.** *RIN3*: deletion on chr14:92611452-92611515, duplication on chr14:92369331-92644481, duplication on chr14:92531510-92573409, deletion on chr14:92535796-92535871, and deletion on chr14:92605335-92607867. **H-I.** *ADAM10*: deletion on chr15:58720431-58721649, deletion on chr15:58621731-58622007. **J.** Deletion on *CASP7*: chr10:113725274-113726288.

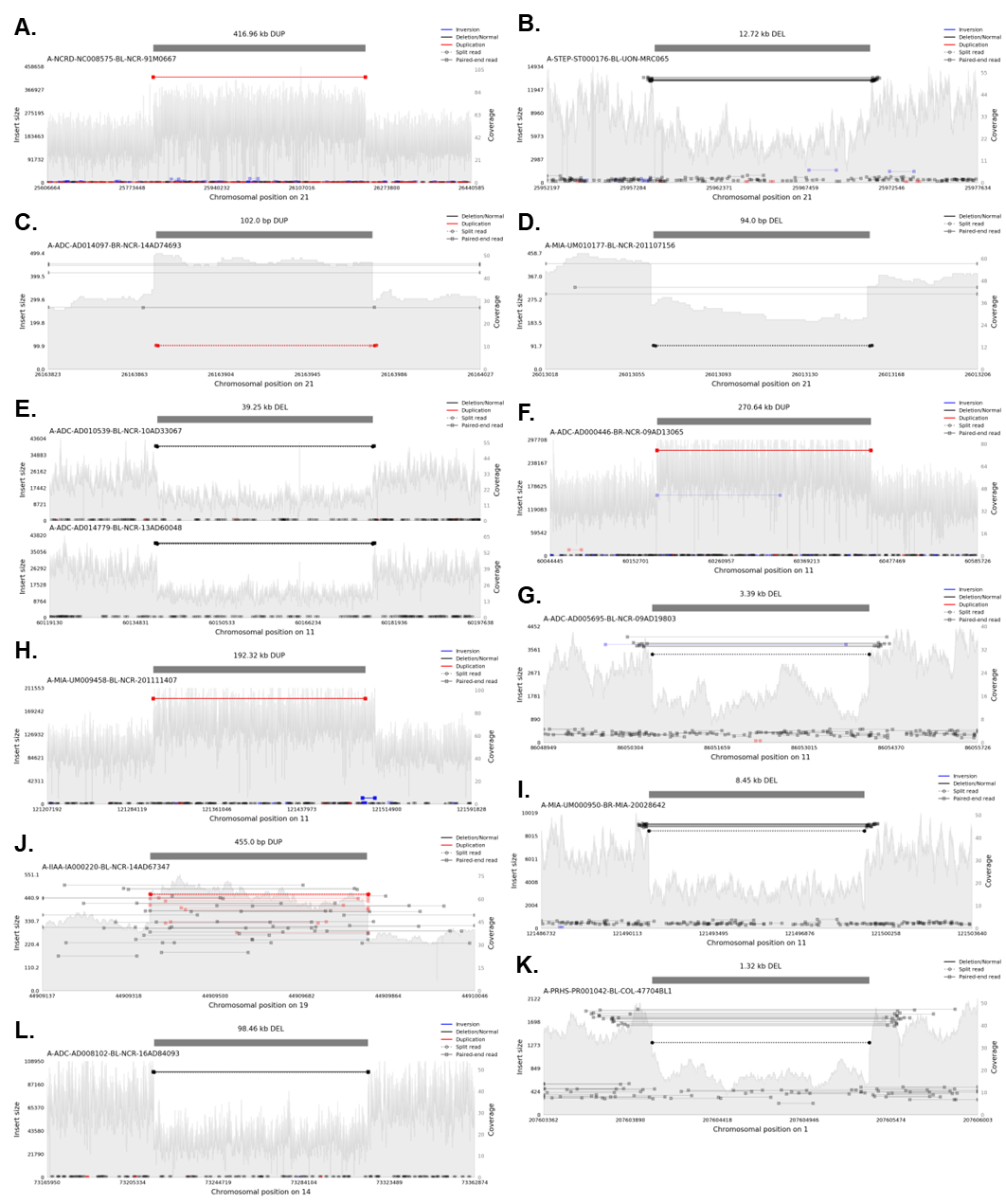

Figure S22. Ultra-rare CNVs on APP, MS4A6A, PICALM, SORL1, APOE, CR1 and PSEN1

**A-D.** *APP*: duplication on chr21:25815144-26232105, deletion on chr21:25958556-25971275, duplication on chr21:26163874-26163976, and deletion on chr21:26013065-26013159. **E-F.** *MS4A6A*: deletion on chr11:60138757-60178011 and duplication on chr11:60179765-60450406. **G.** Deletion on *PICALM*: chr11:86050643-86054032. **H-I.** *SORL1*: duplication on chr11:121303351-121495669, deletion on chr11:121490959-121499413. **J.** Duplication on *APOE*: chr19:44909364-44909819. **K.** Deletion on *CR1*: chr1:207604022-207605343. **L.** Deletion on *PSEN1*: chr14:73215181-73313643.

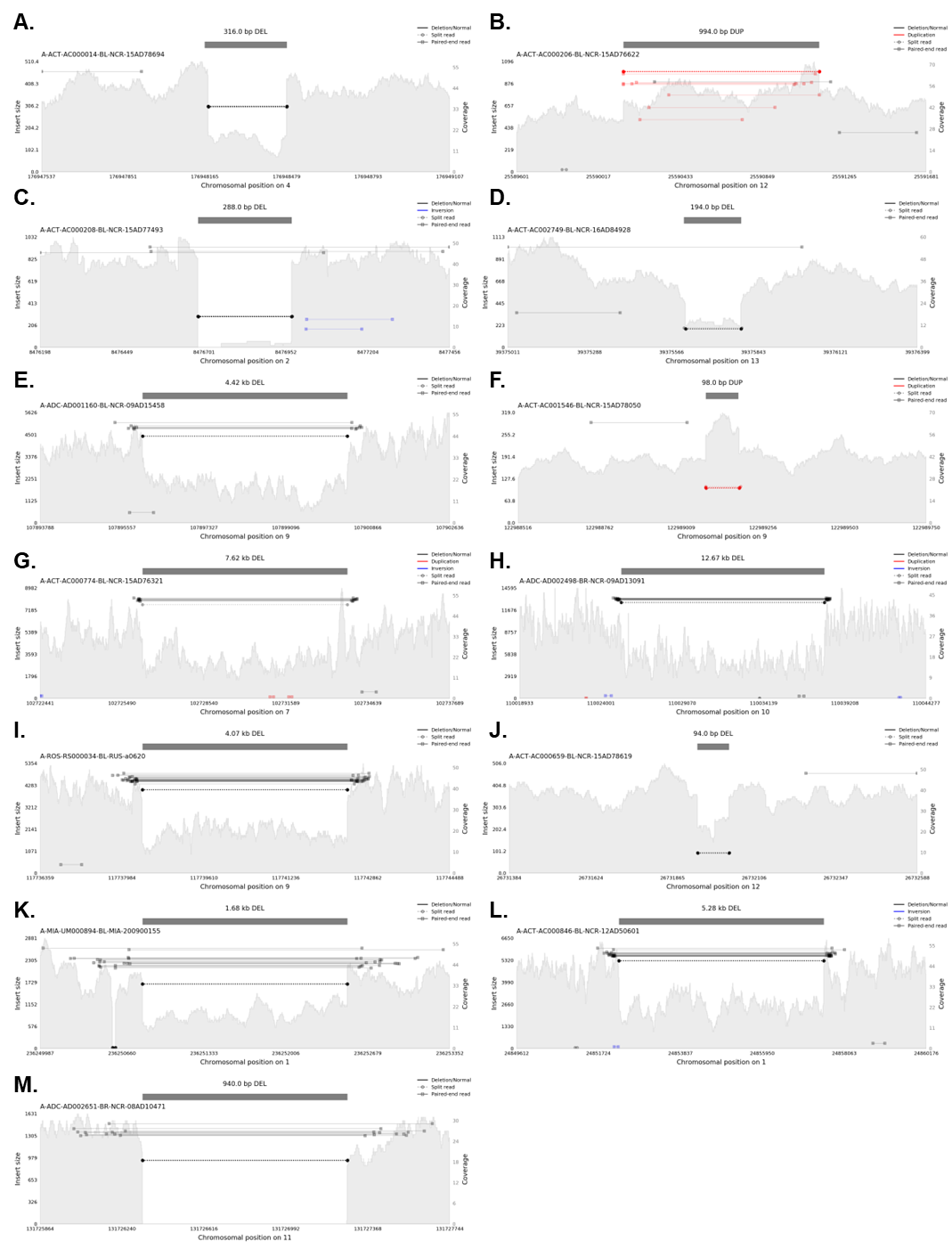

Figure S23. Samplot for deletions and duplications in Table 4

**A-M** represents Samplot for one selected sample for chr4:176948164-176948480:DEL, chr12:25590144-25591138:DUP, chr2:8476683-8476971:DEL, chr13:39375608-39375802:DEL, chr9:107896000-107900424:DEL, chr9:122989084-122989182:DUP, chr7:102726253-102733877:DEL, chr10:110025269-110037941:DEL, chr9:117738391-117742456:DEL, chr12:26731939-26732033:DEL, chr1:236250828-236252511:DEL, chr1:24852253-24857535:DEL, chr11:131726334-131727274:DEL, respectively.

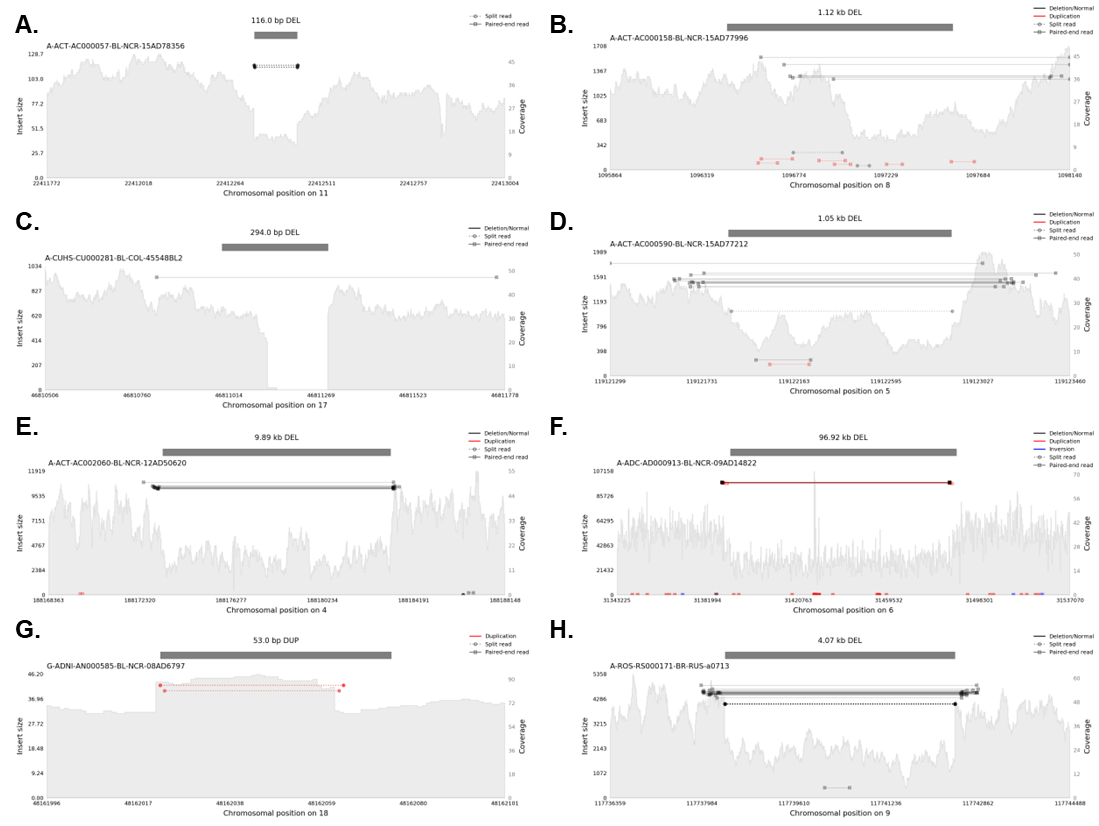

Figure S24. Samplot for deletions and duplications in Table 5

**A-H** represents Samplot for one selected sample for chr11:22412330-22412446:DEL, chr8:1096443-1097561:DEL, chr17:46810995-46811289:DEL, chr5:119121855-119122904:DEL, chr4:188173309-188183202:DEL, chr6:31391686-31488609:DEL, chr18:48162022-48162075:DUP, chr9:117738391-117742456:DEL, respectively.
